# Patterns of opioid use after surgical discharge: a multicentre, prospective cohort study in 25 countries

**DOI:** 10.1101/2023.09.30.23296378

**Authors:** TASMAN Collaborative, Chris Varghese

## Abstract

**Background:** Excessive post-surgical opioid prescribing is contributing to the growing opioid crisis. Prescribing practices are modifiable, yet data to guide appropriate prescription of opioids at surgical discharge remain sparse. We therefore aimed to evaluate the factors associated with opioid consumption following discharge from surgery.

**Methods:** We performed an international, prospective, multicentre, cohort study between 4 April 2022 and 4 September 2022 among adult patients undergoing common general, orthopaedic, gynaecological and urological operations, with follow-up 7 days after hospital discharge. The primary outcome measure was the quantity of prescribed and consumed opioids in oral morphine equivalents (OMEs). Descriptive and multivariable analyses were performed to investigate factors associated with OME quantities prescribed and consumed.

**Findings:** This analysis includes 4273 patients across 144 hospitals in 25 countries. Overall, 30.7% (n=1311) of patients were prescribed opioids at discharge. For those prescribed opioids, a median of 100 OMEs (IQR 60 - 200) were prescribed but only a median of 40 OMEs (IQR 7.5 - 100; p<0.001) were consumed at follow-up 7 days after discharge. After risk-adjustment, an increased amount of opioids prescribed was independently associated with increased opioid consumption in the follow up period (β = 0.33, 95% CI 0.31 - 0.34, p<0.001), and side-effects. The risk of prescribing more opioids than patients’ consumed increased as quantities of opioids prescribed exceeded 100 OMEs, independent of patient comorbidity, procedure, and pain.

**Interpretation:** Patients were prescribed more than twice the quantity of opioids they consumed in the 7 days following discharge from surgery. Prescription quantity was associated with increased consumption of opioids even after adjusting for pain levels, suggesting that prescribing practice is a modifiable risk factor to curtailing excessive opioid consumption. Current quantities of opioids provided are in excess of patient needs and may contribute to increasing community opioid use and circulation.

**Funding:** Maurice and Phyllis Paykel Trust, Surgical Research Funds University of Newcastle.

**Research in context:** *Evidence before this study:* Opioids are frequently prescribed at discharge after surgery, yet little is understood about the drivers of opioid use in this setting. We conducted a literature search between November 2020 and February 2021 for studies reporting on opioid prescription and consumption after discharge from surgery. We used the search terms “opioid”, “surgery”, “discharge”, and applied no language or date restrictions. Several global studies examined variations in opioid prescribing, however, little data exists specific to surgical practice. Several single centre and retrospective surgical series examined the independent role of prescribing practice on opioid consumption; however, these data are not globally generalisable. A recent systematic review and meta-analysis suggests the analgesic efficacy of opioids in the post-surgical-discharge setting may be overstated, exposing populations to their adverse events with minimal improvements in pain management. Given the lack of global, generalisable, high-quality data in the setting of post-surgical discharge, practice is predominantly guided by clinician preferences, dogma, and health system cultures.

*Added value of this study:* This prospective, international, cohort study provides high-quality, cross-specialty, patient-reported data after surgical discharge following a variety of common surgical procedures, including both emergency and elective, minor and major, surgeries. This study includes 4273 patients from 144 centres across 25 countries. Among those prescribed opioids, the median prescription of opioids was 100 oral morphine equivalents (OMEs; IQR 60 - 200) and median consumption at 7-days follow-up was 40 OME (IQR 7.5 - 100; p<0.001). Prescription and consumption of opioids varied by specialty, but predominantly prescribed quantities were in excess of what was consumed by patients within the first 7 days after hospital discharge. This was particularly evident for patients prescribed over 100 OMEs. The quantity of opioids prescribed was associated with higher patient-reported opioid consumption at surgical discharge, and increasing quantities of opioids prescribed and consumed were associated with increased risk of opioid-related harm.

*Implications of all the available evidence:* Overprescribing opioids increases absolute consumption of opioids, even after adjusting for patients’ pain levels, with an associated increase in opioid-related side effects. The value of opioids after surgical discharge has been questioned, and when prescribed, are frequently in excessive quantities. Prescribing practices need to be altered with a more cautious approach to prescribing opioids after surgical procedures. When required, quantities should be rationalised to minimise opioid-related harm, community circulation of opioids, dependence, misuse, and overdose. Our study bridges a crucial knowledge gap and offers guidance on opioid prescribing across a range of common surgical procedures.

## Introduction

The opioid epidemic is a major public health crisis. The age-standardised prevalence of opioid dependence has been estimated to be 510 per 100,000, with the highest prevalence in the United States of America (USA), the Middle East, and East Asia.^1^ This translated to approximately 109,500 opioid overdose deaths worldwide in 2017.^1^ Post-surgical opioid prescribing is a significant contributor to the global opioid crisis,^2^ with overprescribing being an ongoing source of community diversion of unused opioids, misuse, abuse and dependence.^3, 4^

Opioid analgesia, though commonly prescribed to manage postoperative pain, entails a significant potential for harm,^5^ and is therefore facing increasing global scrutiny. A recent systematic review of 47 randomised trials found that opioid prescribing at surgical discharge following minor and moderate elective surgeries did not reduce patients’ pain intensity but did increase adverse events such as vomiting.^6^ Amidst growing awareness of the contribution of excessive and unsafe opioid prescribing to the current opioid crisis,^7–11^ further data are urgently required to guide their clinical use following surgery. As the second highest prescribers of opioids, surgical teams are an important target group for improving prescribing practices.^2^

The Opioid PrEscRiptions and Usage After Surgery (OPERAS) study therefore aimed to quantify the current global practice of opioid prescribing and consumption patterns after discharge from common surgical procedures, and identify factors associated with increased opioid consumption.

## Methods

### Ethical approval

Ethical approvals were obtained according to the requirements at each participating centre and verified by the central steering committee. The Hunter New England Human Research Ethics Committee (2021/ETH11508) approved the protocol as the lead site.

### Study design

This was an international, prospective, multicentre, observational cohort study. Analyses were based on a prespecified, published, protocol and the study was registered in the Australian New Zealand Clinical Trials Registry (ANZCTR: ACTRN12621001451897p).^12^ This study is reported in accordance with the STROBE guidelines.^13^

All hospitals routinely performing general, orthopaedic, gynaecological and urological procedures were eligible to enrol. Prospective data was collected from inpatient clinical records and a standardised patient telephone interview undertaken at 7 days post-discharge.^14^ Data collection took place over six predefined 14-day data collection periods between 4 April 2022 and 4 September 2022. Centres could choose to participate in multiple 14-day consecutive recruitment periods.

### Eligibility criteria

Participating centres prospectively screened and approached all consecutive patients that met eligibility criteria to obtain informed participant consent where this was a requirement of the site ethics approval. Participants could withdraw at any stage. Consecutive adult patients aged ≥18 years undergoing either elective (planned) or emergency (unplanned) common general surgical (cholecystectomy, appendicectomy, inguinal hernia repair, colon resection, fundoplication, or sleeve gastrectomy), orthopaedic (total or reverse shoulder arthroplasty, rotator cuff or labral repair, anterior cruciate ligament repair, or hip or knee arthroplasty), gynaecological (hysterectomy, oophorectomy, or salpingectomy and oophorectomy), and urological procedures (prostatectomy, cystectomy or nephrectomy) were eligible to be included.^12^ Only patients discharged home or to a non-healthcare setting were included. Patients receiving medication-assisted treatment of opioid dependence with methadone, suboxone, or buprenorphine, discharged to rehabilitation, nursing supported care services, another hospital, or discharged with palliative intent were excluded. Patients undergoing multivisceral resections or who required return to theatre were also excluded.

### Outcome and explanatory variables

The primary outcome was the proportion of prescribed opiates that were consumed within 7-days post-discharge.^15, 16^ This is in line with guideline-based recommendations for duration of post-surgical discharge opioid prescriptions.^16, 17^ Data were also collected on patient demographics (age, gender, tobacco use, vaping status, alcohol use, BMI, Society of Anesthesiologists (ASA) physical status classification), comorbidities, diagnosis and procedure specific details (indication, surgical approach, and urgency), opioid use in the 24 hours prior to hospital discharge, opioid prescription at the time of discharge from hospital (including opioid type, dose, and quantity of pills), patient reported outcomes (including patient-reported opioid consumption including type, dose, and quantity of pills), post-operative complications; and requirement for further analgesia. Data on opioid doses were converted to oral morphine equivalents (OME) to account for the potencies of different medications and allow comparison. OME conversion ratios were calculated using conversion ratios defined by the Australian and New Zealand College of Anaesthetics (ANZCA) Faculty of Pain Medicine.^18^ Where opioid conversion ratios were not defined by ANZCA, accepted conversion ratios were identified through a literature search and agreed upon by consensus from members of the OPERAS Scientific Advisory Group.^12^ Methods for calculation of OMEs are further detailed in **Table S1**. Cumulative OME doses were used to enable pragmatic comparisons; irrespective of intended duration of prescriptions this represents the quantity of opioids provided to patients. Opioid side effects were defined as one or more of the following: nausea or vomiting, drowsiness, itching, dizziness, or constipation at the 7-day follow-up phone call. Demographics and opioid prescribing practices were compared between high income countries (HIC) and low and middle income countries (LMIC) as defined by Organisation for Economic Co-operation and Development (OECD).^19^

### Statistical analysis

All statistical analyses were performed with R version 4.2.0 (R Foundation for Statistical Computing, Vienna, Austria) using the *tidyverse*, *rms* and *finalfit* packages. An *a priori* sample size calculation was performed, necessitating a minimum sample size of 852.^12^

Factors collected for patients lost to follow-up (but not those who withdrew consent) were compared with the included cohort to assess any selection bias in those lost to follow-up. Missing data were explored via visual inspection. The *mice* package was used to perform multiple imputation by chained equations for ASA grade, alcohol consumption, and BMI categories which were assumed to be missing at random, and imputed models were pooled per Rubin’s rules.^20^

Descriptive statistics were used to compare demographic and prescription-specific variables based on whether or not patients were prescribed opioids at discharge using the χ2 test for categorical variables and the Kruskal-Wallis test for continuous variables. The univariable correlation between quantity of opioid in OME prescribed and consumed at 7-days follow-up was depicted using a generalised additive model. The risk of opioid-related side effects was modelled using binomial logistic regression and the independent variables; total OMEs prescribed at discharge, and total OMEs consumed at follow-up were plotted with a spline term. Factors associated with the quantity of opioids prescribed and consumed were modelled using separate mixed-effects hierarchical linear regression with the country, and hospital as the random effect. The model predicting quantity consumed was bootstrapped and applied at the patient-level to quantify with 95% confidence intervals for the adjusted rate of overprescription. Residual, Q-Q plots, and variance inflation factors were interrogated to assess model assumptions.

Multivariable binary logistic regression models for the risk of overprescription (defined as prescribed OME quantity exceeding consumed OME quantity at follow-up) were generated. Sensitivity analyses for various thresholds of overprescription, including prescribed OME quantity exceeding 25%, 50% and 100% of consumed OME quantity, were also performed. Thereafter, the multivariable risk-adjusted odds ratio for overprescription was plotted against the quantity of OME prescribed at discharge, quantity of OMEs consumed 24 hours prior to discharge, and severity of pain experienced in the week after discharge, each with a spline term. Covariate selection for adjusted analyses was considered *a priori* and guided by clinical plausibility, AIC criteria, and model parisomony.^12^ A two-tailed α level was set at 5% for interpretation of significance.

## Results

Between 4 April 2022 and 4 September 2022, data from 4273 patients across 144 hospitals in 25 countries were collected and analysed (2271 women, 53.1%; median age 50 years; **Figure 1** and **Table 1 and S7**). Some 1923 (45.0%) patients were recruited from high-income countries, and 2350 (55.0%) from low- and middle-income countries (**Table S2**). A third of patients were prescribed opioid analgesia at discharge after one of 19 eligible surgical procedures (30.7% [n=1311] overall; **Table 1**). Patients were followed up at a median of 7 days (IQR 7 - 8).

**Figure 1:**
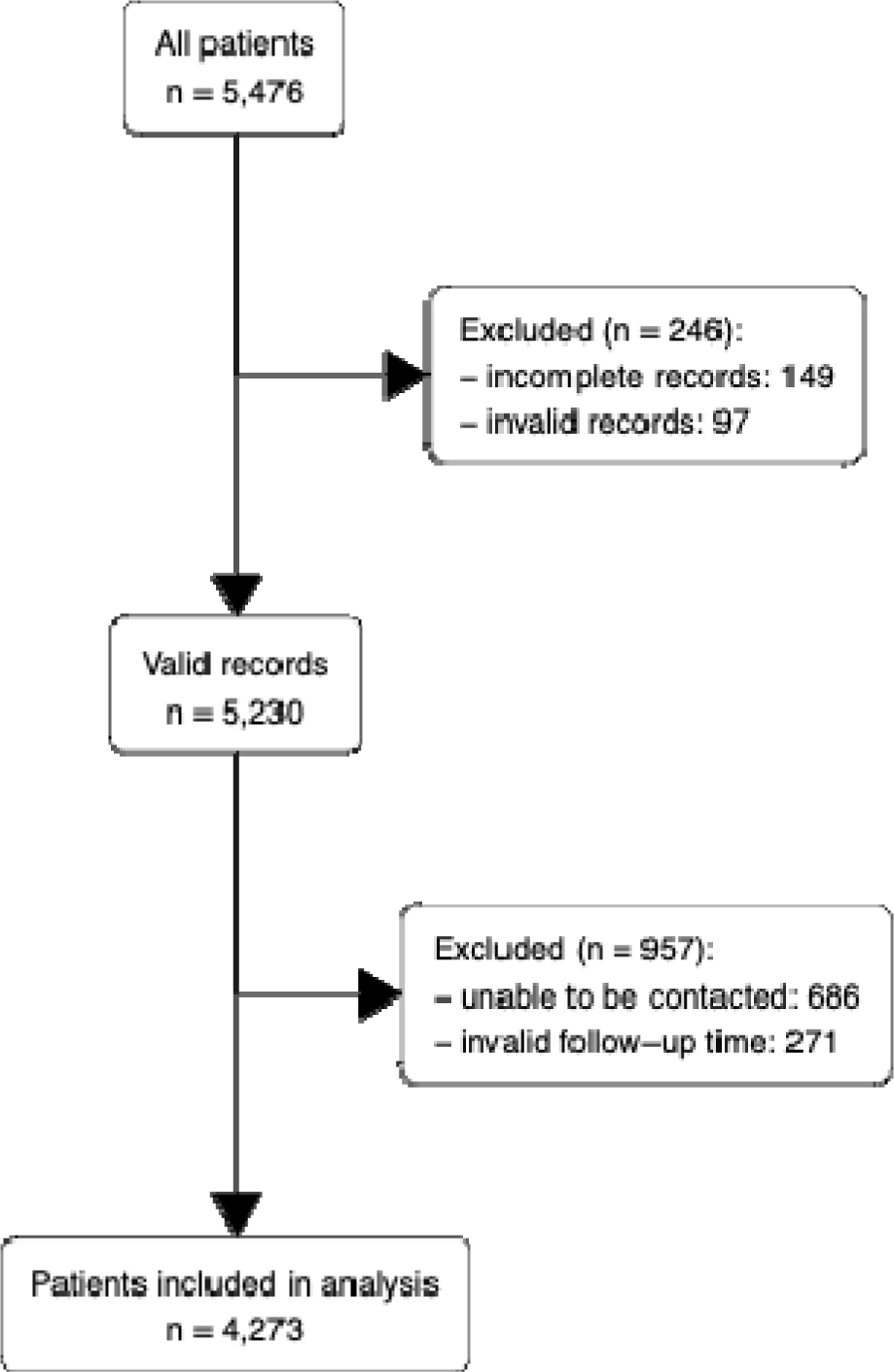
Patients included in analysis and reasons for exclusion Comparison of pre-discharge factors such as age, gender, comorbidity, indication, and specialty showed similar proportions across these factors between those lost to follow-up and those included in analysis (**Table S7**)

**Table 1:**
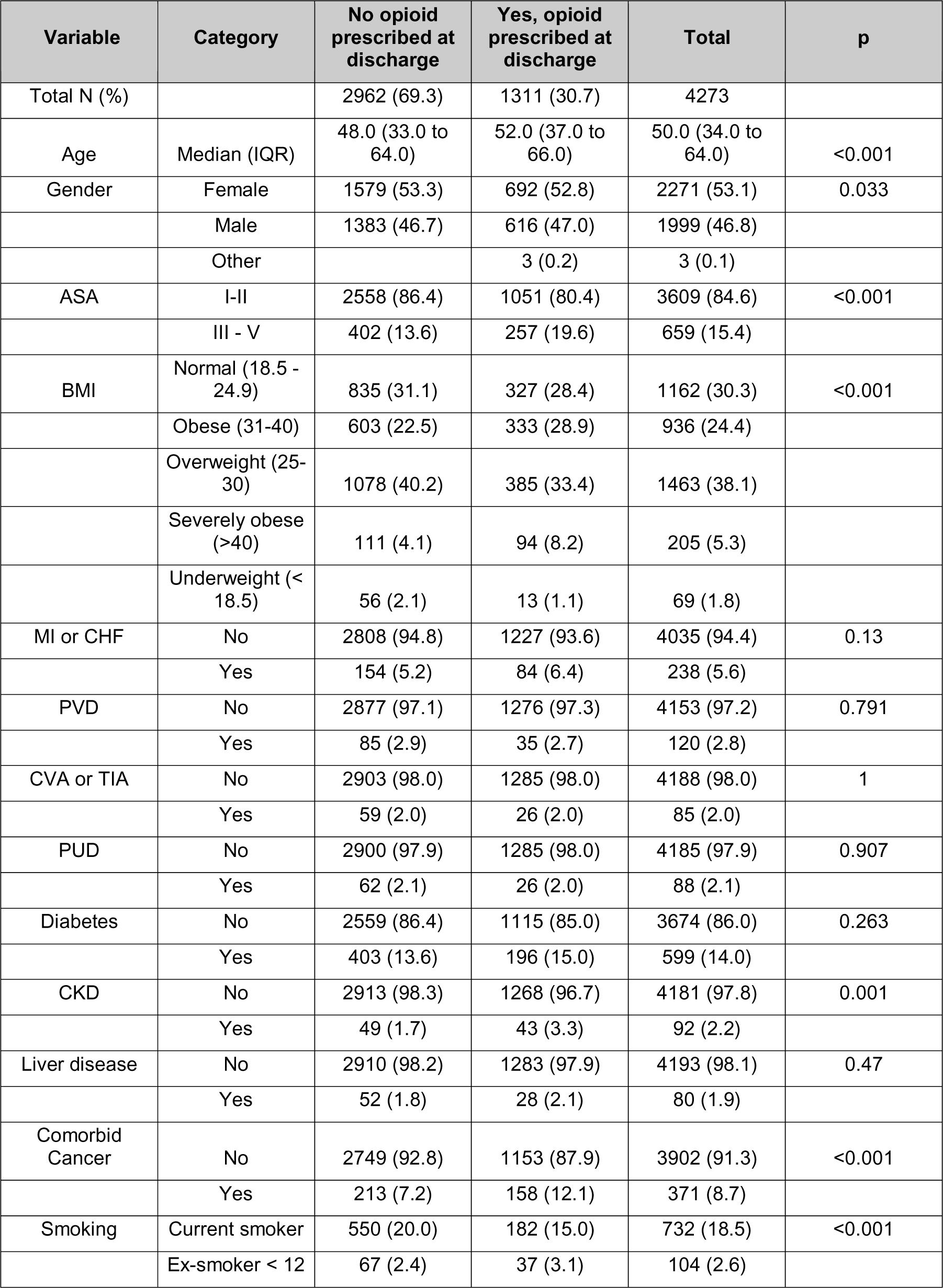
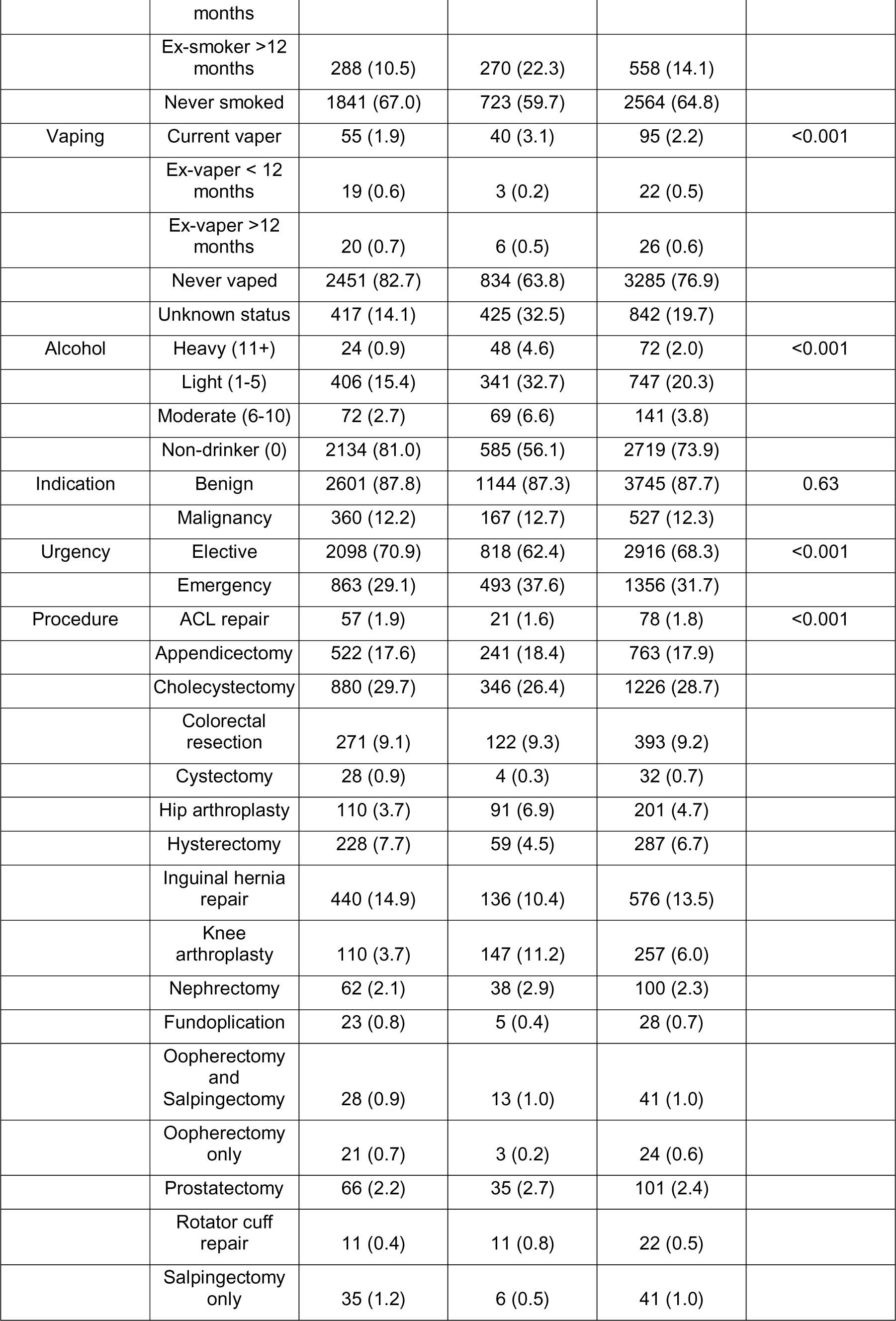
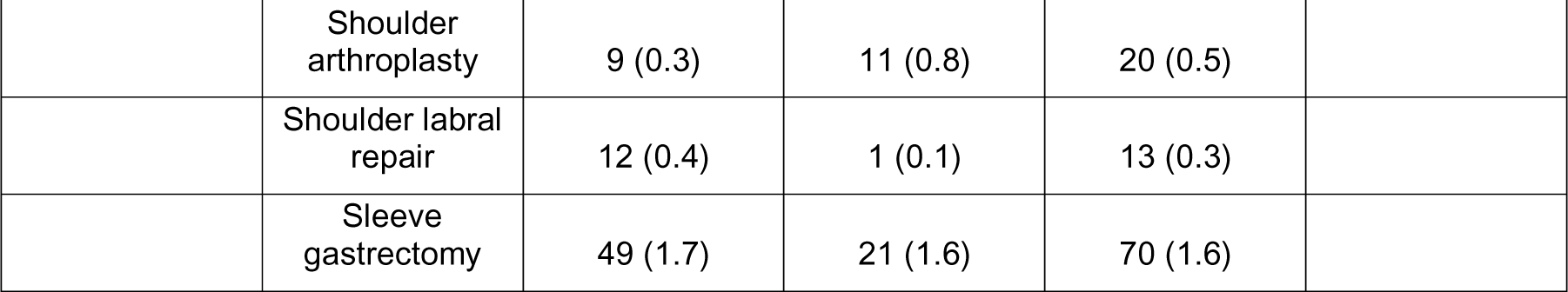
Demographic comparison by opioid prescription at discharge.

### Opioid prescriptions at surgical discharge

Patients who received opioids at discharge tended to be slightly older, with a higher ASA, increased BMI, cancer or kidney disease, smoke or vape, and consume more alcohol (p<0.05; **Table 1**). After risk-adjustment, age (β = −0.30, 95% CI −0.57 - 0.03, p = 0.031), specialty of surgery (β >20, p<0.02) and total OME consumed 24 hours prior to discharge (β 0.6, 95% CI 0.1 - 0.21, p<0.001) were associated with increased quantity of opioid prescribed at discharge (**Table S3**).

Notably, patients were likely to be prescribed significantly more opioid after orthopaedic (β = 89.12, 95% CI 75.29 - 102.94, p<0.001) and gynaecological procedures (β = 20.11, 95% CI 3.47 - 36.75, p = 0.018), compared to general surgical procedures. Of note, following all procedures except arthroplasty, fewer than 50% of patients were prescribed an opioid at discharge (**Figure 2A**). The total amount of opioids consumed 24 hours prior to discharge was also positively associated with larger opioid prescriptions at discharge (β = 0.16, 95% CI 0.1 - 0.21, p<0.001). This mean of the imputed models for the quantity of opioids prescribed had a good fit with conditional R^2^ = 0.39 and the pooled model is summarised in **Table S3**. Importantly, of 1952 patients who received no opioids prior to discharge, 197 (10.1%) received an opioid prescription on discharge.

**Figure 2:**
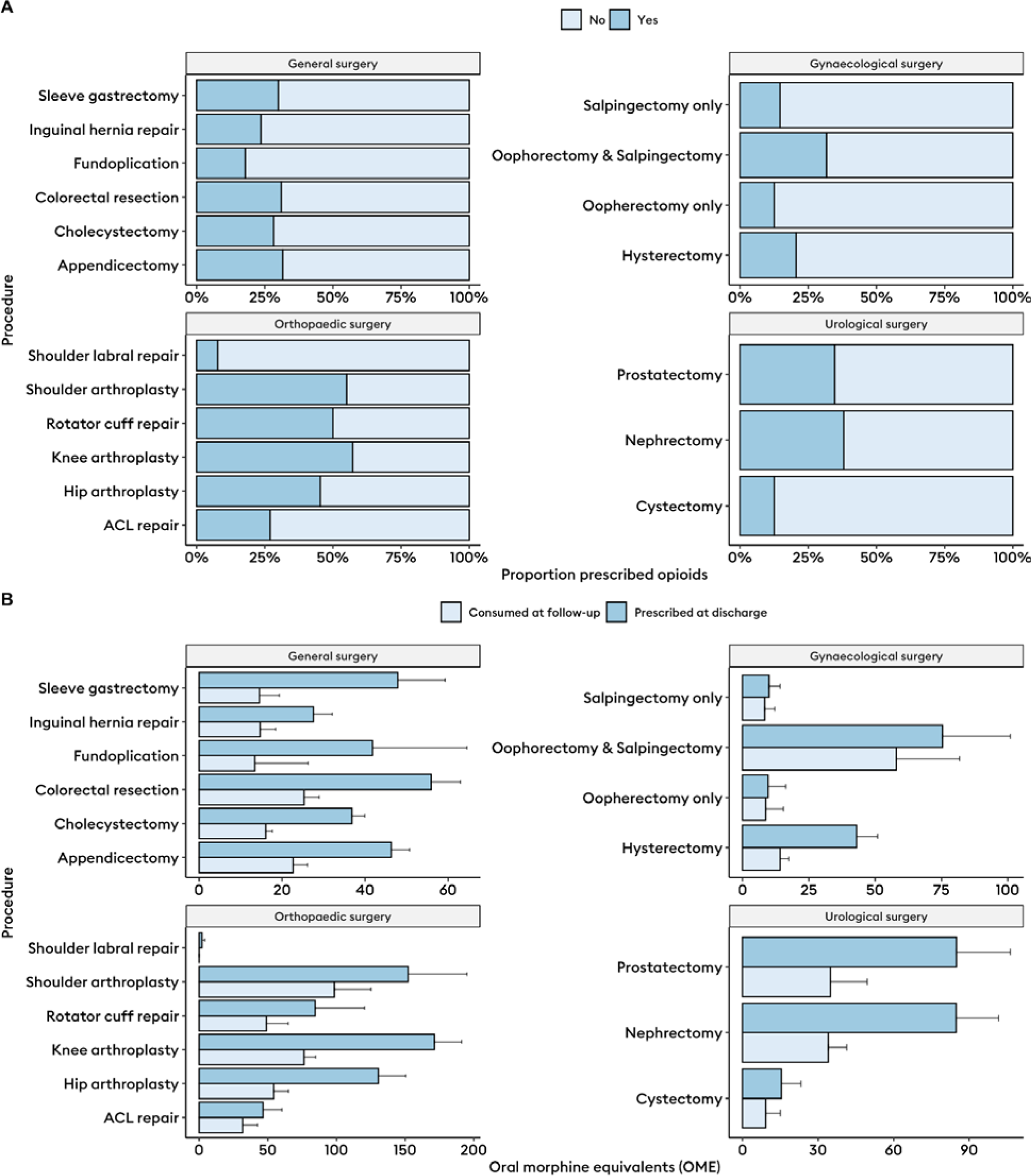
A) Proportion of patients prescribed an opioid at discharge stratified by specialty and surgical procedure (n = 4273). B) Mean and standard error for oral morphine equivalents of opioid prescribed at discharge after surgery and consumed within 7-days follow-up stratified by specialty and surgical procedure (n = 4273). ACL; anterior cruciate ligament.

Patients prescribed opioids at discharge tended to have longer operations (p<0.001), more complications (p<0.001), and were more frequently referred to an acute pain service (8.2% vs 4.7%, p<0.001), but had similar lengths of stay (median 2 days (IQR 1-3) vs 2 days (IQR 1-3), p=0.698) to those not prescribed opioids. Regarding co-analgesics, patients prescribed opioids were more often discharged with paracetamol (89.4% vs 71.4%, p<0.001), gabapentinoids (4.6% vs 1.3%, p<0.001), and tricyclic antidepressants (2.4% vs 0.5%, p<0.001). In the 7-days post-discharge, patients prescribed opioids experienced more pain (median visual analogue rating 20 (IQR 5-40) vs 10 (IQR 0-30), p<0.001). These data are summarised in **Table 2**.

**Table 2:** Clinical and analgesic outcomes by opioid prescription at discharge.

| Variable | Levels | No opioid prescribed at discharge | Yes opioid prescribed at discharge | Total | p |
| --- | --- | --- | --- | --- | --- |
| Total N (%) |  | 2962 (69.3) | 1311 (30.7) | 4273 |  |
| Pain severity | Median (IQR) | 10.0 (0.0 to 30.0) | 20.0 (5.0 to 40.0) | 10.0 (1.0 to 30.0) | <0.001 |
| Operative duration (min) | Median (IQR) | 80.0 (55.0 to 120.0) | 98.0 (66.0 to 135.0) | 87.0 (60.0 to 120.0) | <0.001 |
| Postoperative complications | I | 380 (12.8) | 218 (16.6) | 598 (14.0) | <0.001 |
|  | II | 107 (3.6) | 47 (3.6) | 154 (3.6) |  |
|  | IIIa/IIIb/IVa/IVb | 21 (0.7) | 20 (1.5) | 41 (1.0) |  |
|  | None | 2450 (82.8) | 1025 (78.2) | 3475 (81.4) |  |
| Length of stay (days) | Median (IQR) | 2.0 (1.0 to 3.0) | 2.0 (1.0 to 3.0) | 2.0 (1.0 to 3.0) | 0.698 |
| Referral to acute pain service | No | 2823 (95.3) | 1203 (91.8) | 4026 (94.2) | <0.001 |
|  | Yes | 139 (4.7) | 108 (8.2) | 247 (5.8) |  |
| Discharged with paracetamol | No | 847 (28.6) | 139 (10.6) | 986 (23.1) | <0.001 |
|  | Yes | 2113 (71.4) | 1172 (89.4) | 3285 (76.9) |  |
| Discharged with NSAIDs | No | 1502 (50.7) | 687 (52.4) | 2189 (51.3) | 0.321 |
|  | Yes | 1458 (49.3) | 623 (47.6) | 2081 (48.7) |  |
| Discharged with gabapentinoids | No | 2922 (98.7) | 1251 (95.4) | 4173 (97.7) | <0.001 |
|  | Yes | 38 (1.3) | 60 (4.6) | 98 (2.3) |  |
| Discharged with tricyclic antidepressants | No | 2945 (99.5) | 1279 (97.6) | 4224 (98.9) | <0.001 |
|  | Yes | 14 (0.5) | 31 (2.4) | 45 (1.1) |  |

Of those prescribed opioids, the vast majority were given a single opioid (89.5%), and the majority used this prescription (79%). The majority (85%) of these patients also used paracetamol after discharge. Of note only 37% of patients prescribed an opioid were discharged with laxatives, and 22.5% with antiemetics (**Table 3**). Only 30.5% received documented advice regarding safe disposal of unused opioids.

**Table 3:** Prescribing factors of those prescribed an opioid at discharge.

| **Number of opioids prescribed at discharge** | ***n*** |
| --- | --- |
| **1** | 1174 (89.5) |
| **2** | 131 (10.0) |
| **3** | 6 (0.5) |
| **Total OME prescribed, median (IQR)** | 100.0 (60.0 to 200.0) |
| **Total OME consumed, median (IQR)** | 40.0 (7.5 to 100.0) |
| **Paracetamol used** | 1114 (85.0) |
| **NSAID used** | 506 (38.6) |
| **D/C with laxatives** | 485 (37.0) |
| **D/C with antiemetics** | 295 (22.5) |
| **Safety net advice given** | 377 (30.5) |
| **Prescribed opioids used** | 1035 (79.0) |

### Comparison of prescription quantities to consumption quantities

When prescribed, the median quantity of opioids was 100 OME (IQR 60 - 200). In comparison, at 7 days follow-up, the median quantity consumed by patients was 40 OME (IQR 7.5 - 100; p<0.001; **Figure 3**). The average ratio of the quantity of OMEs predicted to be consumed to what was prescribed was 2.22 (95% CI 2.13 - 2.30; **Table S4**). This trend of prescribing opioids in excess of that consumed by 7 days was evident across most procedures (**Figure 2B**). Increasing quantities of opioids prescribed at discharge was associated with a linear increase in risk of opioid side effects (**Figure 4A**).

**Figure 3:**
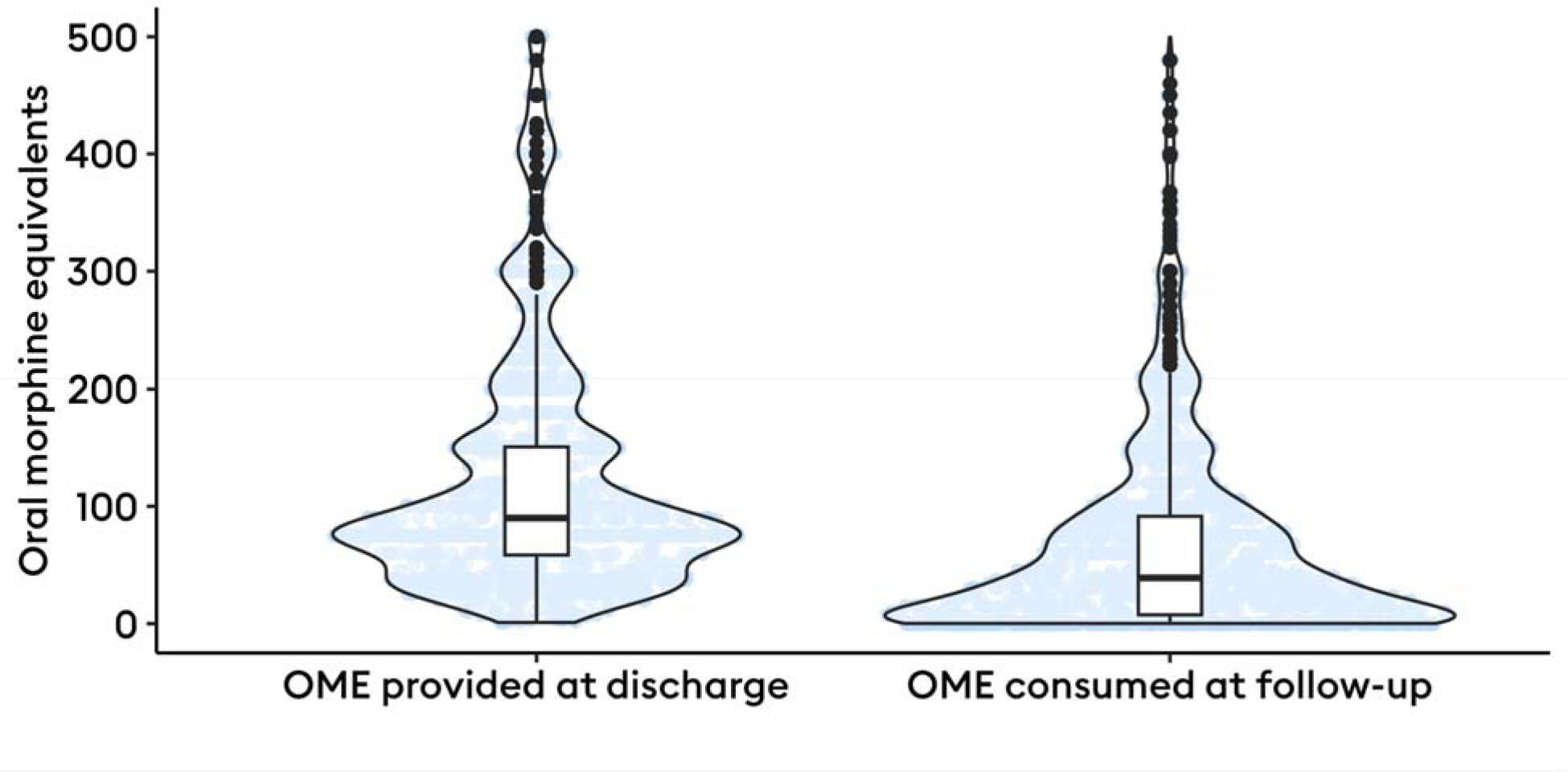
Box and violin plots of total amount of oral morphine equivalents of opioid prescribed at discharge after surgery and consumed within 7-days follow-up

**Figure 4:**
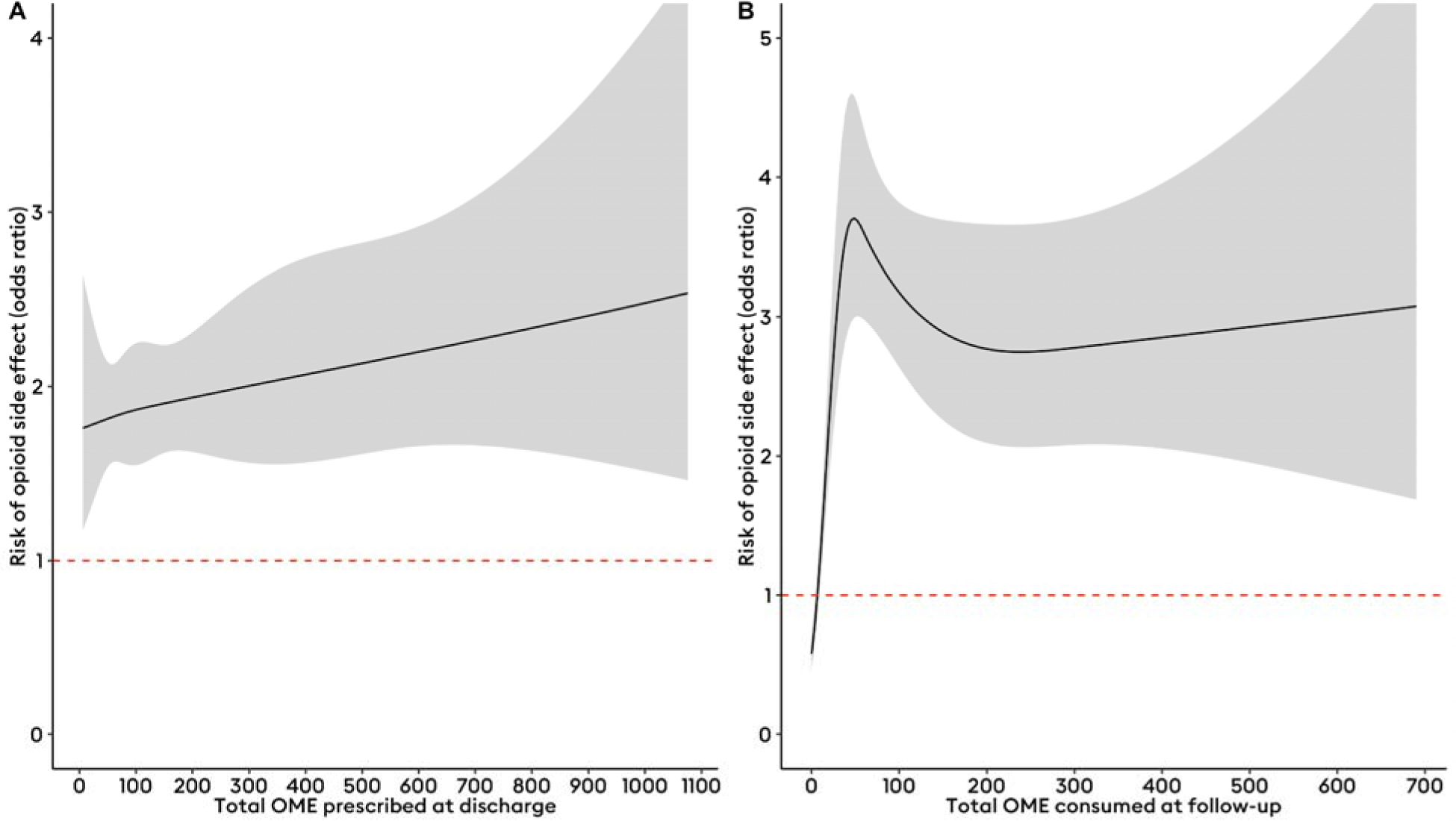
Restricted cubic splines with 4 knots plotting odds ratios of the risk of opioid-related side effects against a spectrum of oral morphine equivalent (OME) totals prescribed at discharge (A) and consumed at follow-up (B). The risk of opioid-related side effects (in odds ratios) is plotted against the total amount of opioids prescribed at discharge and consumed at follow-up (in OMEs) with associated 95% confidence intervals. The red dashed line represents the no-effect line.

Additionally, there was a steep increase in risk of opioid-related side effects with increasing opioid consumption to approximately 50 OMEs, beyond which the risk of side effects was roughly triple that of <10 OMEs (**Figure 4B**). Consumption of opioids at follow-up increased linearly with the quantity of opioids prescribed at discharge (r = 0.57, p<0.001, **Figure S1**). After risk-adjustment, pain severity (β = 0.19, 95% CI 0.10 - 0.27, p<0.001), total amount of opioids prescribed (β = 0.33, 95% CI 0.31 - 0.34, p<0.001), and total amount of opioids consumed 24 hours prior to discharge (β = 0.07, 95% CI 0.04 - 0.1, p<0.001) were independently and positively associated with increased opioid consumption. This mean of the imputed models for the quantity of opioids consumed had a good fit with conditional R^2^ = 0.60 and the pooled model is summarised in **Table S4**.

Overprescription was evident after appendicectomy, inguinal hernia surgery, cholecystectomy, sleeve gastrectomy, fundoplication, colorectal resections, hip arthroplasty, knee arthroplasty, hysterectomy, oophorectomy and salpingectomy, prostatectomy, and nephrectomy (p<0.05; **Figure 2** and **Table 4**). However, 59 (1.4%) patients consumed more opioids than they were initially prescribed (e.g., sources other than the discharge prescription). The risk of overprescribing opioids increases linearly as larger quantities are prescribed at discharge, particularly over 100 OMEs (**Figure 5A**). Below 100 OMEs, the risk of overprescribing progressively reduces (**Figure 5A**). Similarly, consumption of less than 35 OMEs in the 24 hours prior to discharge was predictive of likely overprescription (**Figure 5B**). Risk of overprescription was present irrespective of varying pain levels after discharge (**Figure 5C**). These findings persisted in subgroup analyses where overprescription was defined as prescriptions more than 25%, 50%, and 100% more than what was consumed (**Figure S2**).

**Figure 5:**
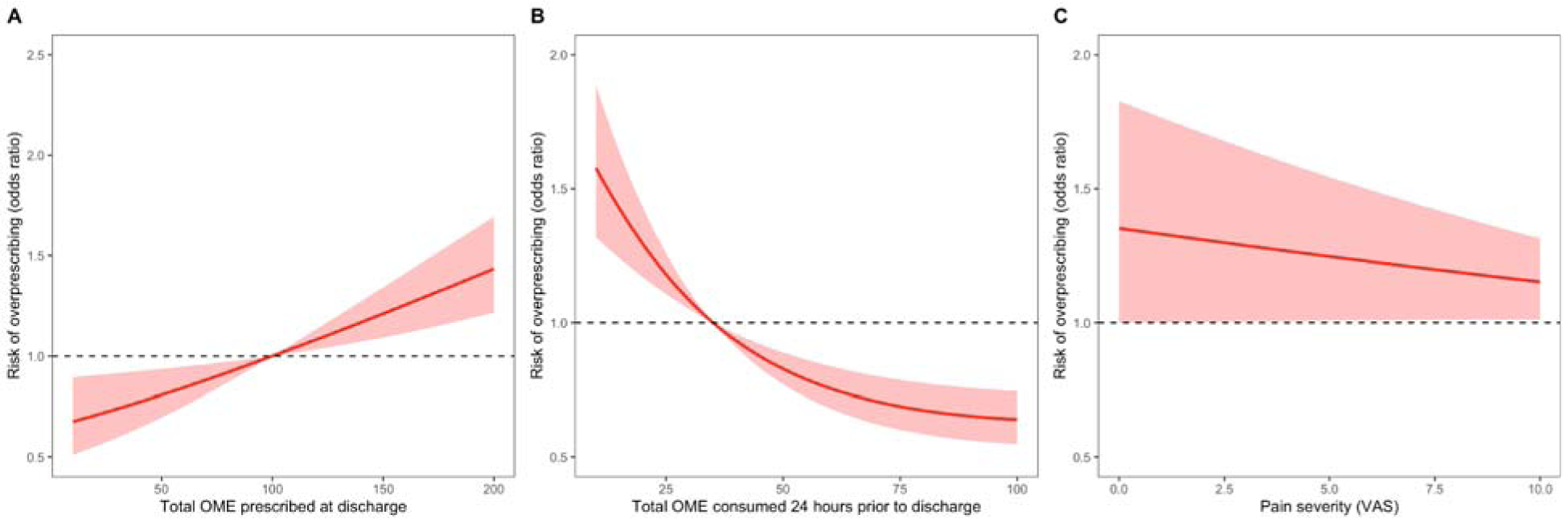
Restricted cubic spline plots with 3 knots for a binary logistic regression model for the risk of over-prescribing opioids (prescribing more than what is consumed at 7-days follow-up). A) Plots risk of overprescription across a spectrum of OMEs prescribed at discharge; B) plots risk of overprescription across a spectrum of OMEs consumed 24 hours prior to discharge; and D) plots risk of overprescription across a spectrum of numeric rating scale pain scores.

**Table 4:** Difference between quantities of opioids prescribed at discharge and consumed at follow-up by procedure; p-values adjusted using Benjamini-Hochberg correction.

| Procedure | n | Mean difference | Adjusted p value | Significance |
| --- | --- | --- | --- | --- |
| ACL repair | 78 | -1.6602635 | 0.101 | ns |
| Appendicectomy | 763 | -5.9886527 | 3.26E-09 | **** |
| Cholecystectomy | 1226 | -8.2838667 | 3.09E-16 | **** |
| Colorectal resection | 393 | -5.5354394 | 5.69E-08 | **** |
| Cystectomy | 32 | -1.2955701 | 0.205 | ns |
| Hip arthroplasty | 201 | -5.5205941 | 1.04E-07 | **** |
| Hysterectomy | 287 | -4.5066117 | 9.61E-06 | **** |
| Inguinal hernia repair | 576 | -5.1066892 | 4.47E-07 | **** |
| Knee arthroplasty | 257 | -6.126131 | 3.38E-09 | **** |
| Nephrectomy | 100 | -3.6260453 | 0.000457 | *** |
| Fundoplication | 28 | -2.0720403 | 0.048 | * |
| Oophorectomy and Salpingectomy | 41 | -2.7921709 | 0.008 | ** |
| Oophorectomy only | 24 | -1 | 0.328 | ns |
| Prostatectomy | 101 | -3.9550139 | 0.000143 | *** |
| Rotator cuff repair | 22 | -1.6152028 | 0.121 | ns |
| Salpingectomy only | 41 | -1.061003 | 0.295 | ns |
| Shoulder arthroplasty | 20 | -1.4645411 | 0.159 | ns |
| Shoulder labral repair | 13 | -1 | 0.337 | ns |
| Sleeve gastrectomy | 70 | -3.0008082 | 0.004 | ** |

### Geographical variation

Overall, 54.5% of patients from HIC as defined by OECD (n = 1923) were prescribed opioids, at a median quantity of 37.5 OME (IQR 0 - 112.5). This compared to only 12.5% of patients from LMIC (n = 2350) with a median quantity of 0 OME (IQR 0 to 0; p<0.001). Median consumption of opioids at 7 days was clinically similar (0 OME [IQR 0 - 0] in LMIC vs 0 OME [IQR 0 - 30] in HIC, p = 0.002). Notably there was significant variation in rates of opioid prescription by hospital centre, but after adjusting for patient factors, clear differences between LMIC and HIC centres were evident (**Figure 8**). These data are summarised in **Table S5, S6** and **Figure 7**.

**Figure 7:**
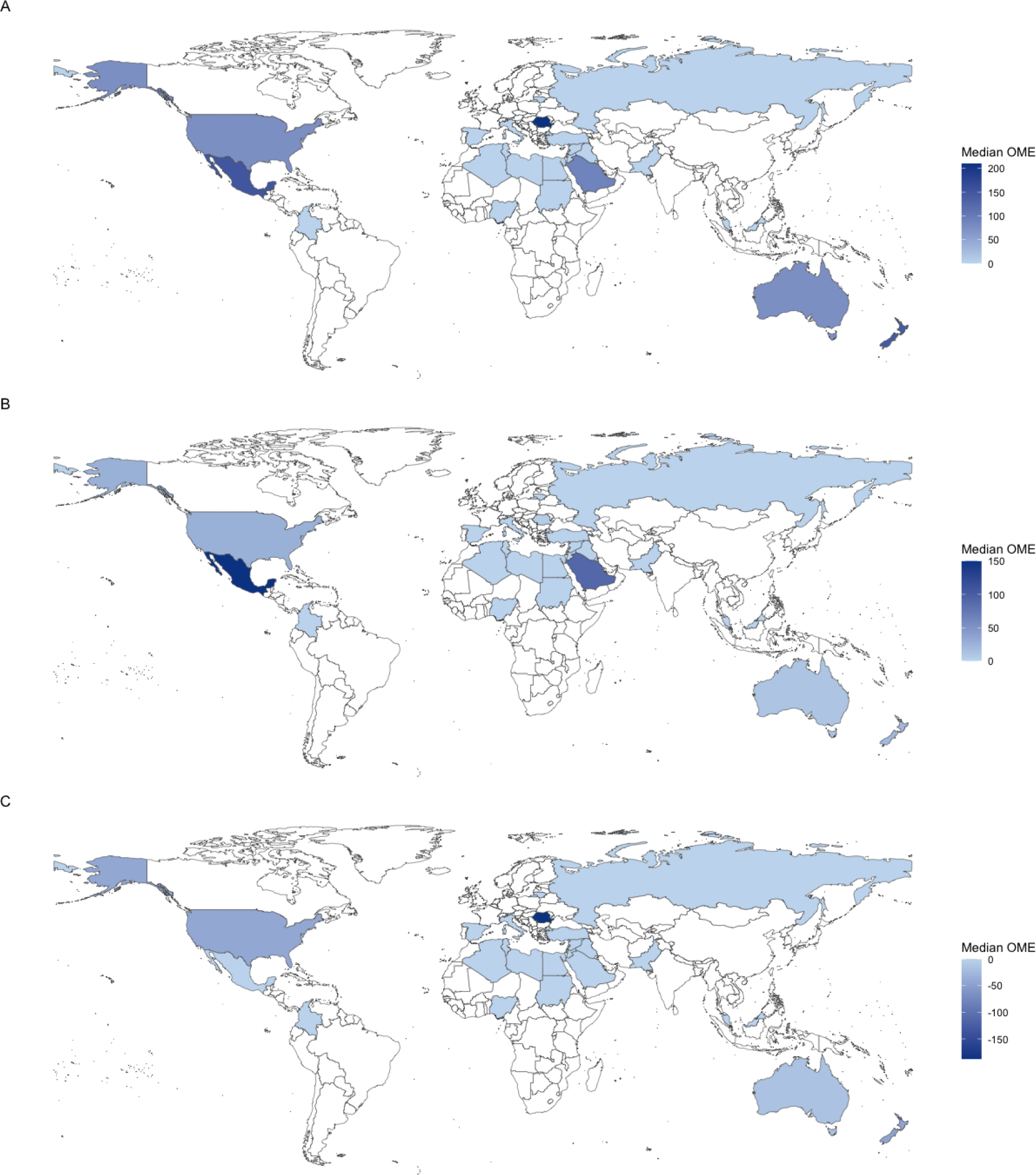
Global variation in A) prescription, B) consumption, and C) differences in prescription and consumption quantities of opioids in oral morphine equivalents (OME)

**Figure 8:**
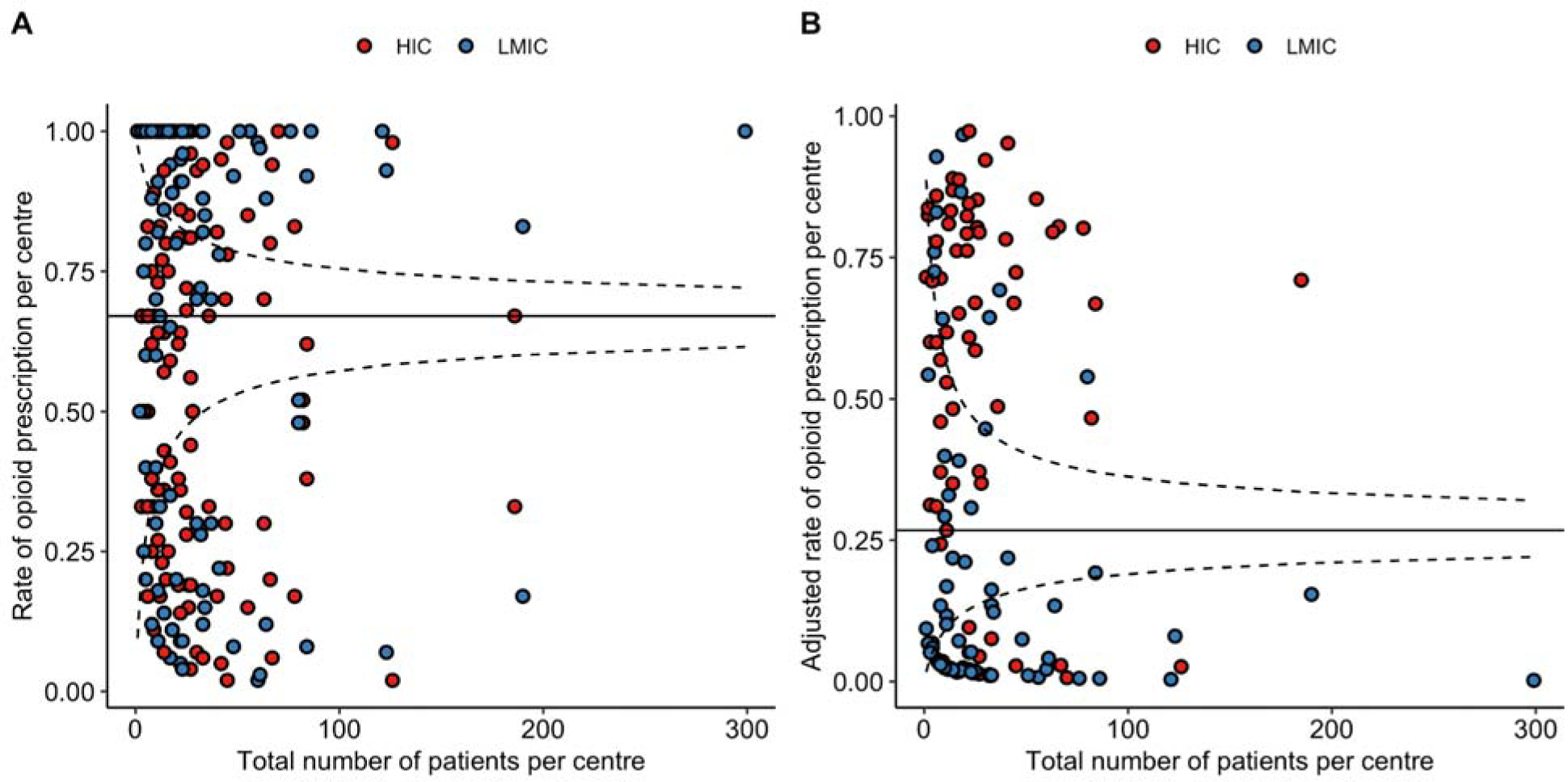
Global variation in rate of opioid prescription by centre, stratified by country income group. A) Unadjusted rates of opioid prescription per centre and B) adjusted rates of opioid prescription per centre (using data from the same model shown in **Table S4**).

## Discussion

In this multinational cohort study opioids were widely prescribed in excess of what patients consumed at 7 day follow up after discharge from common general, urological, gynaecological, and orthopaedic surgical procedures. Prescribing higher quantities of opioids after discharge from surgery was associated with a higher risk of experiencing opioid-related side effects. The quantity of opioids prescribed by clinicians at discharge was associated with increased opioid consumption even after adjusting for post-discharge pain severity and pre-discharge opioid consumption. Excess opioid prescribing was evident across a geographically diverse cohort, particularly in high income countries. These findings confirm that urgent improvements in prescribing practice are needed to mitigate the globally escalating opioid crisis.

Excess opioid prescribing has been described across many surgical specialties.^9–11, 21–25^ Our data corroborate a vast literature predominantly originating from the USA, showing that excess volumes of opioids are prescribed at surgical discharge globally (frequently in excess of 100 OME).^7, 10, 24–26^ We found fewer than 50% of opioids prescribed are actually consumed within 7 days, findings similar to the results of a systematic review of USA studies, which found that only 29-58% of prescribed opioids were consumed post discharge.^9^ This demonstrates that opioid overprescribing at surgical discharge is more widespread than previously accepted. This work also highlights important inequities in global practice, with individuals from HIC being more likely to be prescribed opioids, at higher quantities, compared to patients from LMICs. As efforts are put in place to improve opioid stewardship globally, care must be taken to ensure equitable global prescribing practices at surgical discharge.^27^

Overprescription poses a key risk for increased unregulated circulation of opioids in the community. Safe disposal of excess opioids is known to be low,^28^ and further evidenced by our findings that fewer than one-third of patients received documented advice about safe disposal of opioids. The retention of what is frequently 60% of an individual’s prescription quantity in the community significantly increases the risk of opioid misuse. Lipari *et al* have shown that peers and family remain a much more widespread source of opioids for non-medical use in the community than the black market or “doctor shopping” strategies.^29^ This highlights the responsibility that falls on clinicians to ensure appropriate prescription quantities.

When prescribing opioids after surgery, clinical care standards emphasise a patient-centred approach limiting the duration of usual discharge opioid prescriptions to less than 7 days of short-acting opioids for acute pain,^17, 30^ and this is consistent with the most recent published international multidisciplinary consensus statement on the prevention of opioid-related harm in adult surgical patients.^31^ Providing large quantities of opioids for longer durations poses a substantially increased risk for chronic use, misuse, and overdose.^15^ The duration of the first opioid analgesic prescription has been found to be more strongly related to misuse in the early postoperative period than the dosage, with each refill and week of opioid analgesic prescription associated with 20% increase in opioid misuse among opioid-nailve patients.^32^ Ongoing pain management in the community beyond the first post discharge week should involve the transfer of care to primary healthcare professionals, who are well positioned to ensure appropriate review and to implement weaning plans as appropriate.^17^ Additionally, guideline-based strategies to optimise analgesia at surgical discharge should include using non-opioid analgesia as first-line and utilising multimodal analgesia.^31, 33^ Though we found paracetamol was co-prescribed with opioids in close to 90% of patients discharged with opioids, NSAIDs were co-prescribed in only 50%. Concerningly, 10% of patients not requiring opioids prior to discharge were discharged with opioid analgesia. As previously reported, this variability suggests prescribing practices remain dogmatic, habit-driven, and are in urgent need of reform.^11, 34^

We also show that a key driver of excess opioid prescribing and the subsequent harms are driven by prescriber choices. The quantity of opioids prescribed at discharge is associated with the quantity of opioids consumed, even after adjusting for pre-discharge opioid consumption and post-discharge pain levels. Numerous regional series have identified this trend,^7, 35–37^ and we verify the persistence of such trends in an international multi-specialty cohort. Our data demonstrate the risk of overprescription increases significantly beyond 100 OMEs, even when overprescription was defined as double the quantity of opioids that patients consumed. Our data offers a useful quantitative guide for prescribing. Firstly, as fewer than 50% of patients were prescribed opioids at discharge in this cohort, the decision to prescribe opioids at all should be individualised. Secondly, care should be taken prior to prescribing opioids in excess of the average consumed quantity for each respective procedure (as visualised in **Figure 2**). And finally, particular care should be taken when prescribing in excess of 100 OMEs, as the risk of overprescription increases significantly beyond this point.

Strategies suggested to curtail the volume of opioid prescriptions at surgical discharge in the literature include defining patient needs using surveys, generating operation-specific guidelines, and statistical models to predict patient needs, all of which have been trialled without increases in opioid refills.^26, 36, 38^ It is widely accepted that opioid prescribing should be individualised, however reliably determining patient-needs remains challenging. In the setting of plastic surgery, Zhang *et al* proposed a predictive model for postoperative discharge opioid requirements based on inpatient opioid consumption trends, and whose accuracy is not influenced by age, gender identity, procedure type, length of stay, or preoperative opioid use.^37^ While this was derived from a small cohort of plastic surgery patients, such a data-driven approach seems a promising intervention to guide opioid prescribing. Building a predictive model for consumption that is generalisable to a diverse range of patients irrespective of geography and procedure is the focus of ongoing work with the OPERAS dataset.

This is a global, prospective, multi-specialty study assessing opioid prescriptions and patient-reported opioid consumption. We had high levels of data-completion and minimal loss to follow-up (<20%) for a study requiring telephone interviews of patients at such a scale. Nevertheless, there are several limitations to our study, including need for care interpreting causality owing to the observational nature of the data. In addition, elements of subjectivity and recall bias are inevitable with patient-reported data points, but are mitigated by the short, 7-day follow-up time point after discharge. Guidelines recommend no longer than a week’s supply of opioids should be prescribed after surgery,^16, 17^ to encourage patients with inadequately managed pain to seek help and to mitigate large opioid prescription volumes. Hence, our 7-day follow-up is both clinically and pragmatically optimal.^15, 16^ Further, we do not explore long-term clinical outcomes and dependence beyond patient-reported pain in the acute post-discharge setting.This data also amalgamates a geographically diverse cohort where opioid prescribing practices vary, but this is also a strength of the study that adds to the generalisability of the findings.

Our findings have direct implications for clinical practice, highlighting the importance of appropriate post-discharge opioid prescribing to reduce opioid-related harm, such as excess diversion of unused opioids into the community, dependence, misuse, and overdose. As suggested by Howard *et al*, our multi-centre data can define “consumption norms” to generate procedure-specific guidelines for widespread use;^7^ these should then be disseminated through professional bodies such as ‘NPS MedicineWise’ to improve clinical practice. Targeting prescribing education and change interventions at early career prescribers, who frequently organise post-surgical discharges, will be pivotal.

Opioid prescribing after surgery is a global issue, with significant implications for patients. We prescribe nearly double the quantity of opioids patients consume in the post-discharge period, exposing them to opioid-related harm. Individualised opioid prescribing at discharge remains important; however, the quantities currently provided are in excess of patient needs and are driving increased consumption of opioids. While patient pain levels, and pre-discharge opioid consumption influence opioid consumption at discharge, the quantity of opioids prescribed remains a modifiable factor to curtailing excessive prescriptions of unused opioids.

## Data Availability

All data produced in the present study are available upon reasonable request to the authors contingent on ethical approval(s).

## SUPPLEMENTARY MATERIAL

### Supplementary Methods

Variations to prespecified protocol:

- Other aims stated in the protocol, including impact of opioid prescription and consumption on patient-reported outcomes and quality of life, are reported in a separate manuscript.

### OPERAS Opioid prescription and consumption calculations

#### OPERAS Opioid Calculations

The following values were calculated for OPERAS data analysis.

A. Opioid consumption in 24 hours prior to discharge (as oral morphine equivalents (OME))
B. Total quantity of opioids prescribed on discharge (as OME)
C. Opioid consumption in 7 days after discharge (as OME)

##### A. Opioid consumption in 24 hours prior to discharge (as OME)

###### Calculation

For each opioid: Total quantity consumed (mg/mcg) x relevant conversion factor (depending on opioid type and route of administration, see **Table 1**) = OME for that opioid. Sum of all individual OMEs = total opioid consumption in 24 hours prior to discharge (as OME).

##### B. Total quantity of opioids prescribed on discharge (as OME)

###### Calculation

For each opioid: Total quantity of medication prescribed (number of tablets/ patches/ injections/ volume of liquid) x dose (mg/mcg) x relevant conversion factor = OME for that opioid. Sum of all OMEs = total opioid prescription at discharge (as OME).

##### C. Opioid consumption in 7 days after discharge (as OME)

###### Calculation

For each opioid: Total quantity of medication consumed (number of tablets/ patches/ injections/ volume of liquid) x dose (mg/mcg) x relevant conversion factor = OME for that opioid. Sum of all OMEs = total opioid consumption in 24 hours prior to discharge (as OME).

#### Oral Morphine Equivalent (OME) Calculation

In order to calculate the OME for a given opioid, the opioid dose is multiplied by the relevant conversion factor (listed in Table 1).

For example, oral oxycodone can be converted to oral morphine using a conversion factor of 1.5. Therefore, the OME of oxycodone 5mg is 5 x 1.5 = 7.5mg.

OMEs with corresponding ANZCA FPM conversion factors were integrated with the REDCap data collection tool. Other OMEs were calculated using R.

Opioid combination products such as Oxycodone/ Naloxone were coded according to the opioid component.

**Table S1:**
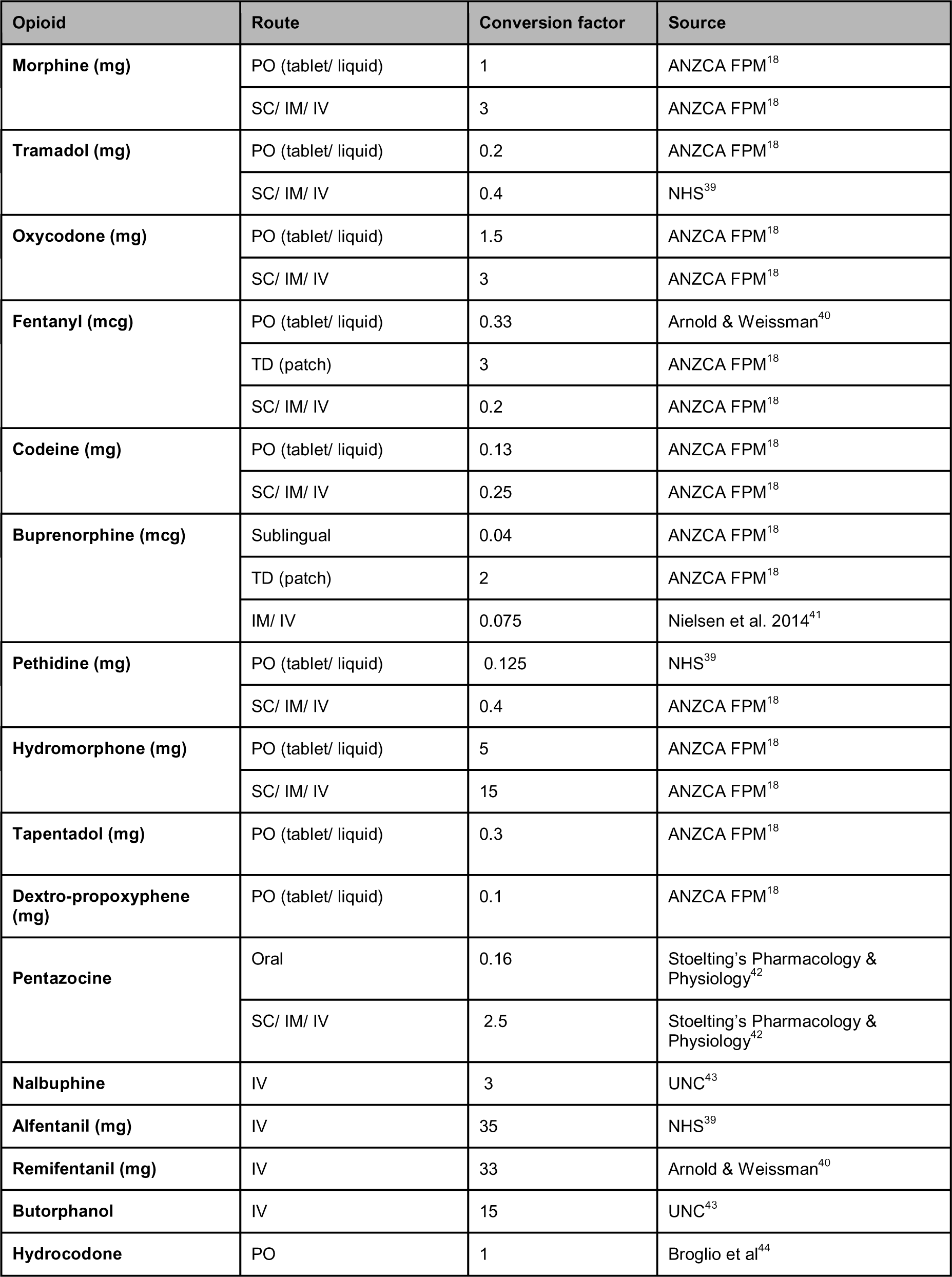
Opioid Oral Morphine Equivalent Conversion Factors used in OPERAS and corresponding source.

| Opioid | Route | Conversion factor | Source |
| --- | --- | --- | --- |
| **Morphine (mg)** | PO (tablet/ liquid) | 1 | ANZCA FPM<sup>18</sup> |
|  | SC/ IM/ IV | 3 | ANZCA FPM<sup>18</sup> |
| **Tramadol (mg)** | PO (tablet/ liquid) | 0.2 | ANZCA FPM<sup>18</sup> |
|  | SC/ IM/ IV | 0.4 | NHS<sup>39</sup> |
| **Oxycodone (mg)** | PO (tablet/ liquid) | 1.5 | ANZCA FPM<sup>18</sup> |
|  | SC/ IM/ IV | 3 | ANZCA FPM<sup>18</sup> |
| **Fentanyl (mcg)** | PO (tablet/ liquid) | 0.33 | Arnold & Weissman<sup>40</sup> |
|  | TD (patch) | 3 | ANZCA FPM<sup>18</sup> |
|  | SC/ IM/ IV | 0.2 | ANZCA FPM<sup>18</sup> |
| **Codeine (mg)** | PO (tablet/ liquid) | 0.13 | ANZCA FPM<sup>18</sup> |
|  | SC/ IM/ IV | 0.25 | ANZCA FPM<sup>18</sup> |
| **Buprenorphine (mcg)** | Sublingual | 0.04 | ANZCA FPM<sup>18</sup> |
|  | TD (patch) | 2 | ANZCA FPM<sup>18</sup> |
|  | IM/ IV | 0.075 | Nielsen et al. 2014<sup>41</sup> |
| **Pethidine (mg)** | PO (tablet/ liquid) | 0.125 | NHS<sup>39</sup> |
|  | SC/ IM/ IV | 0.4 | ANZCA FPM<sup>18</sup> |
| **Hydromorphone (mg)** | PO (tablet/ liquid) | 5 | ANZCA FPM<sup>18</sup> |
|  | SC/ IM/ IV | 15 | ANZCA FPM<sup>18</sup> |
| **Tapentadol (mg)** | PO (tablet/ liquid) | 0.3 | ANZCA FPM<sup>18</sup> |
| **Dextro-propoxyphene (mg)** | PO (tablet/ liquid) | 0.1 | ANZCA FPM<sup>18</sup> |
| **Pentazocine** | Oral | 0.16 | Stoelting's Pharmacology & Physiology<sup>42</sup> |
|  | SC/ IM/ IV | 2.5 | Stoelting's Pharmacology & Physiology<sup>42</sup> |
| **Nalbuphine** | IV | 3 | UNC<sup>43</sup> |
| **Alfentanil (mg)** | IV | 35 | NHS<sup>39</sup> |
| **Remifentanil (mg)** | IV | 33 | Arnold & Weissman<sup>40</sup> |
| **Butorphanol** | IV | 15 | UNC<sup>43</sup> |
| **Hydrocodone** | PO | 1 | Broglio et al<sup>44</sup> |

**Table S2:**
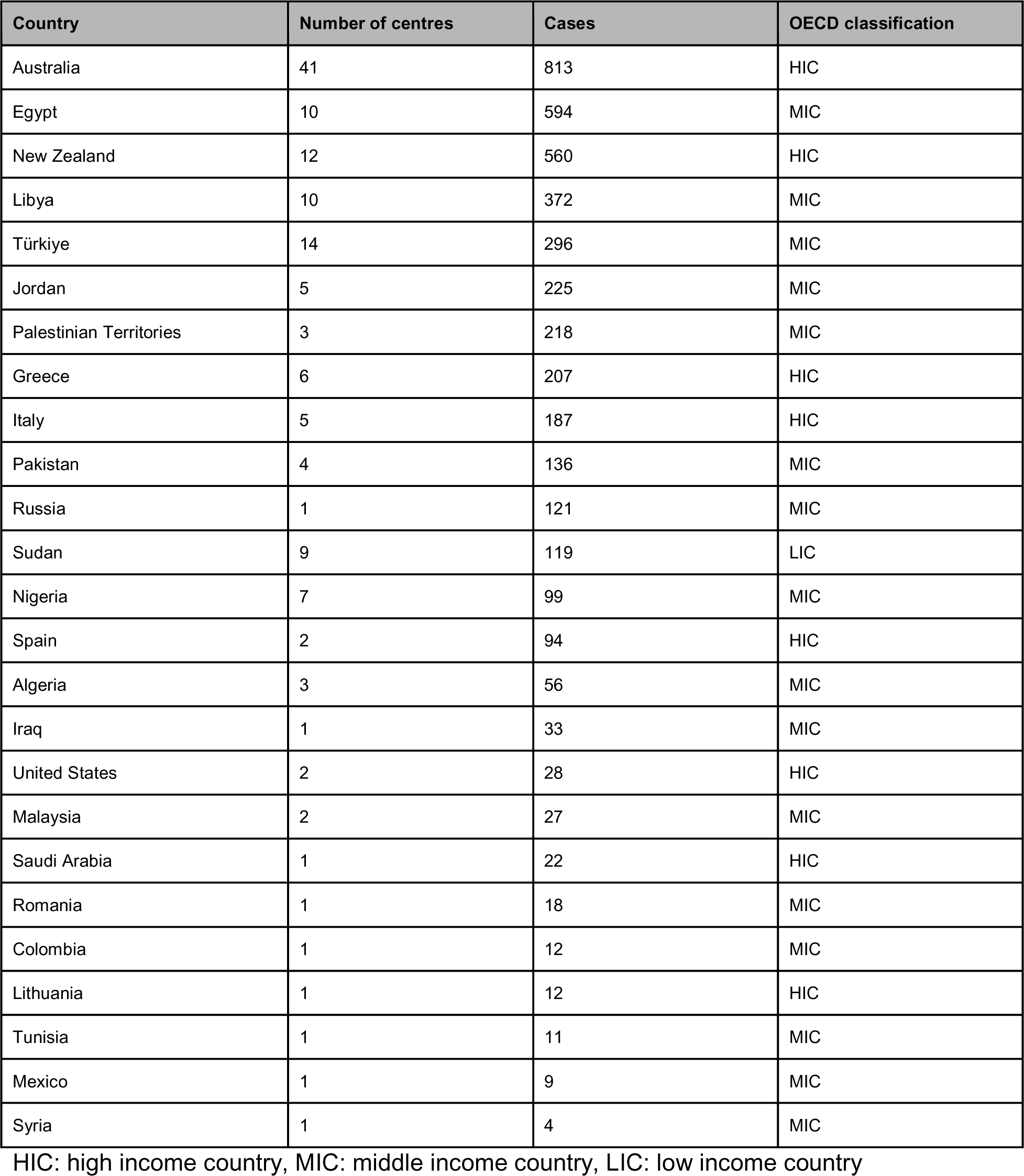
Contributions to study by country.

**Table S3:** Mixed effects hierarchical linear regression model for the quantity of opioids (OME) prescribed to patients at discharge after surgery.

| Factors | $\beta$ coefficient | 95% CI | p value |
| --- | --- | --- | --- |
| Age | -0.3 | -0.57 - -0.03 | 0.031 |
| Gender |  |  |  |
| Male | 2.67 | -6.45 - 11.79 | 0.566 |
| Smoking status |  |  |  |
| Current smoker | -5.84 | -18.34 - 6.67 | 0.361 |
| Ex-smoker < 12 months | -7.89 | -34.58 - 18.8 | 0.563 |
| Ex-smoker >12 months | -3.25 | -17 - 10.51 | 0.644 |
| Alcohol status |  |  |  |
| Heavy (11+) | -1.52 | -40.46 - 37.43 | 0.94 |
| Light (1-5) | 8.55 | -5.8 - 22.9 | 0.246 |
| Moderate (6-10) | 2.17 | -21.05 - 25.4 | 0.855 |
| Indication |  |  |  |
| Malignant | 13.28 | -0.71 - 27.28 | 0.063 |
| BMI |  |  |  |
| Obese (31-40) | 4.39 | -7.25 - 16.02 | 0.46 |
| Overweight (25-30) | 7.84 | -2.22 - 17.91 | 0.127 |
| Severely obese (>40) | 18.45 | -1.34 - 38.25 | 0.068 |
| Underweight (< 18.5) | -1.43 | -35.47 - 32.6 | 0.934 |
| ASA |  |  |  |
| III-V | -4.45 | -16.58 - 7.67 | 0.471 |
| Specialty of operation |  |  |  |
| Gynaecological surgery | 20.11 | 3.47 - 36.75 | 0.018 |
| Orthopaedic surgery | 89.12 | 75.29 - 102.94 | <0.001 |
| Urological surgery | 32.91 | 12.39 - 53.43 | 0.002 |
| Total amount of opioids prescribed 24 hours prior to discharge (OME) | 0.16 | 0.1 - 0.21 | <0.001 |

**Table S4:**
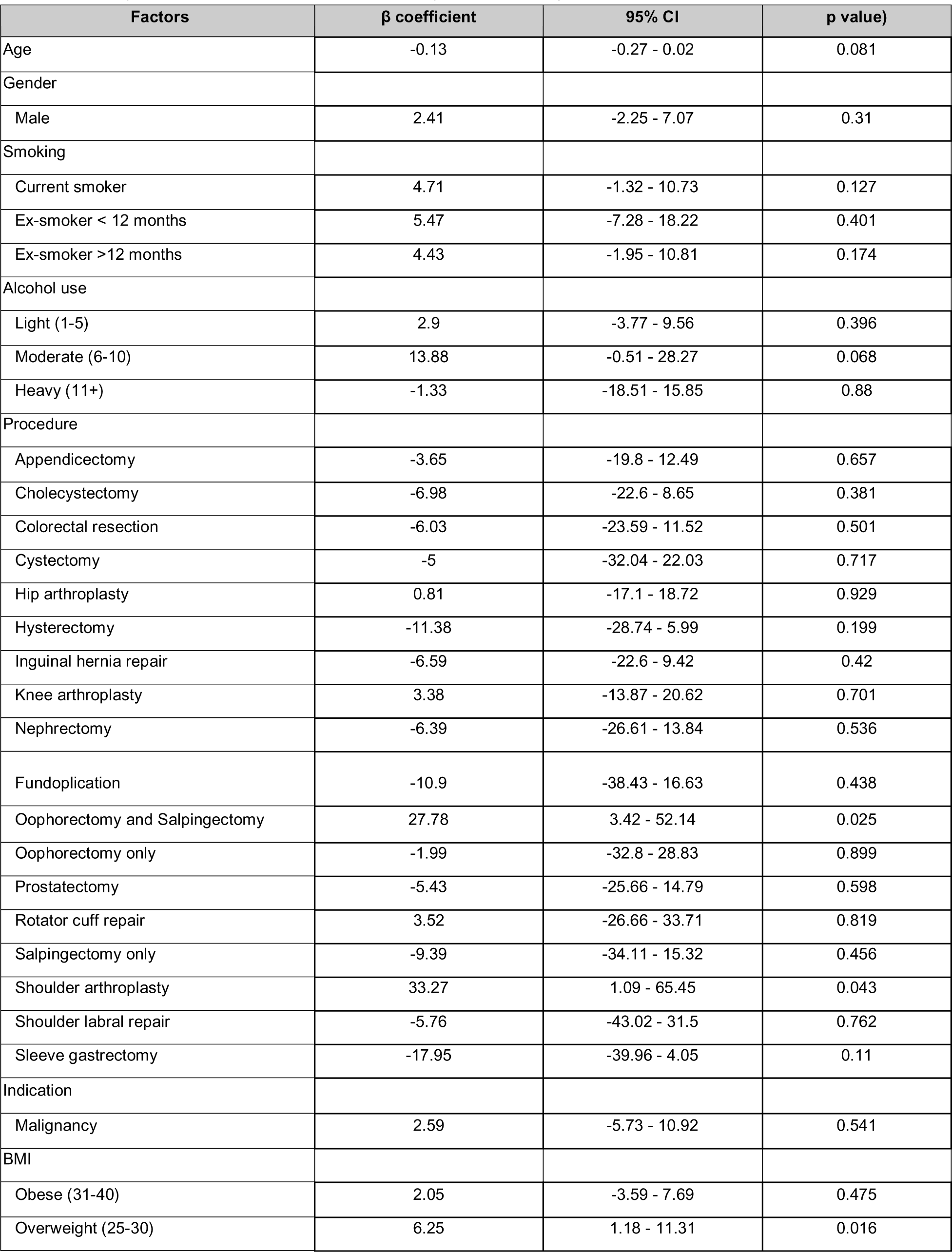
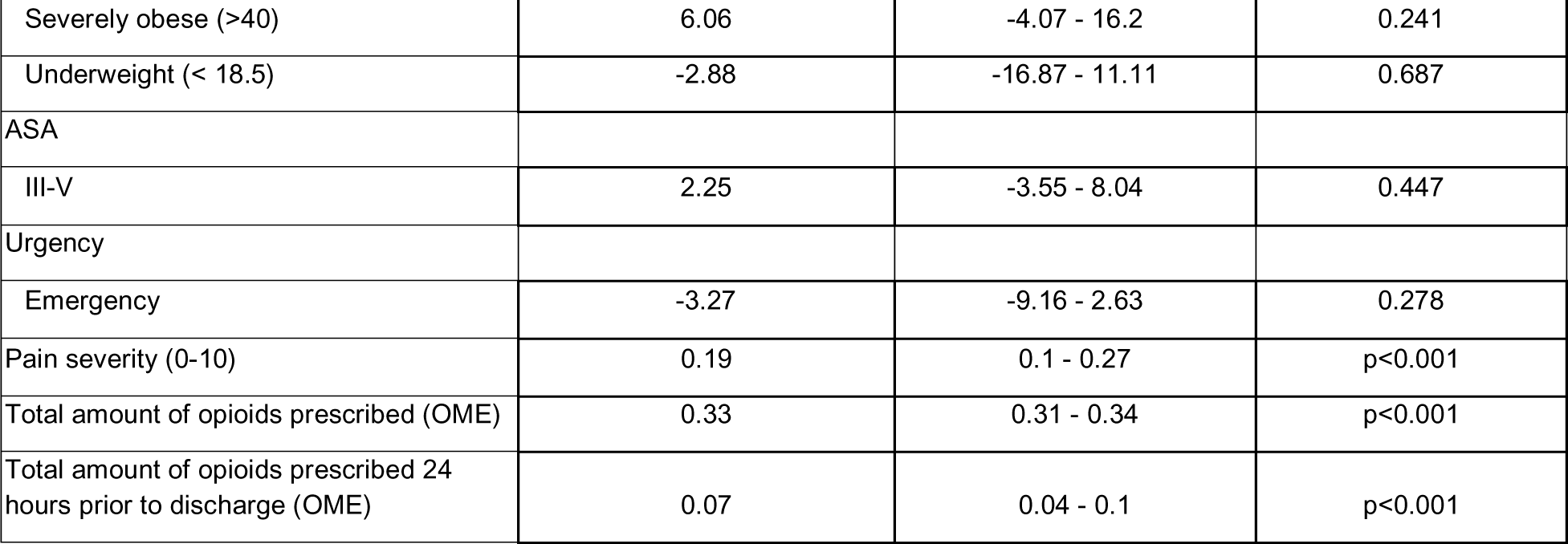
Mixed effects hierarchical linear regression model for the quantity of opioids (OME) consumed by patients at follow-up after surgical discharge.

**Table S5:**
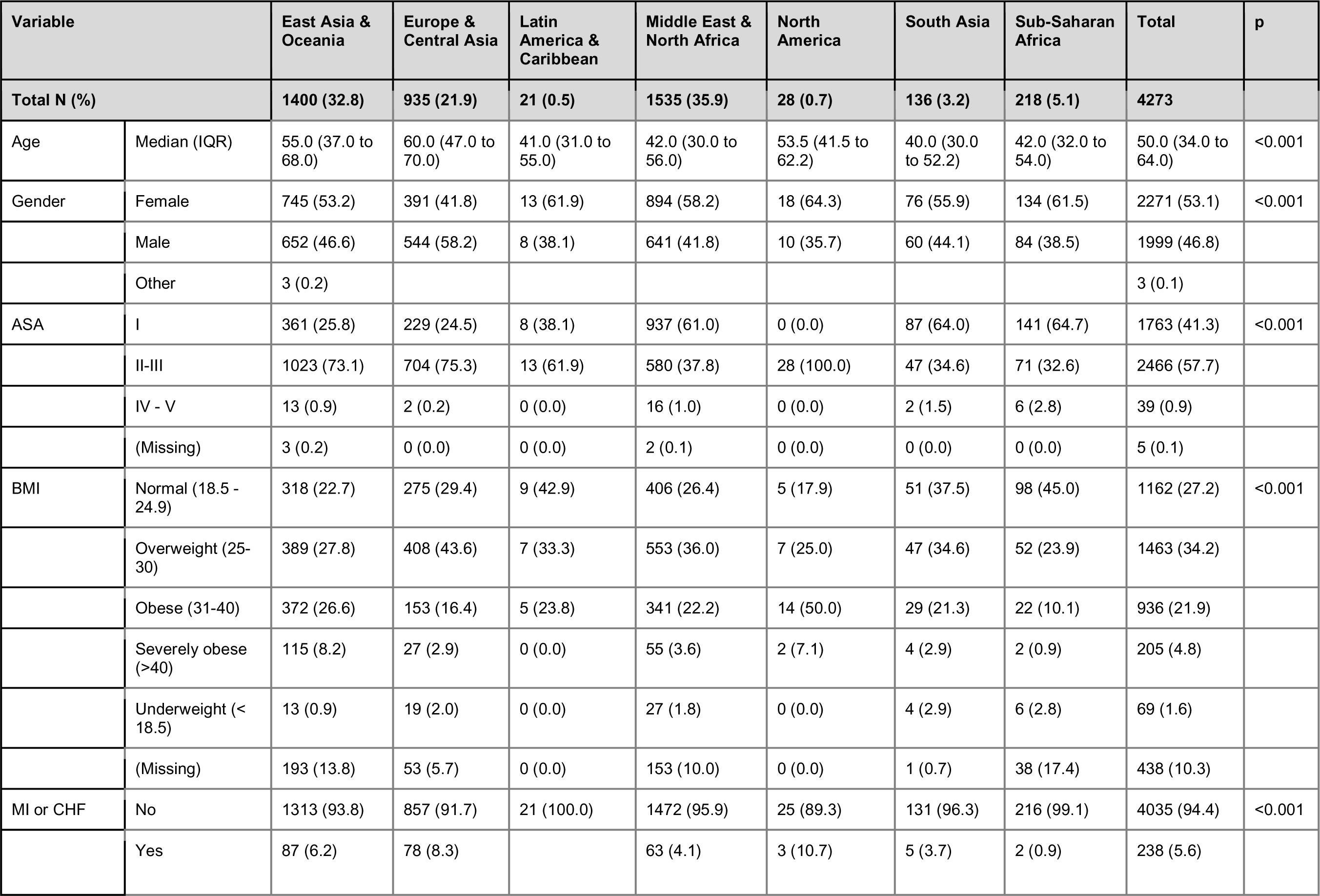
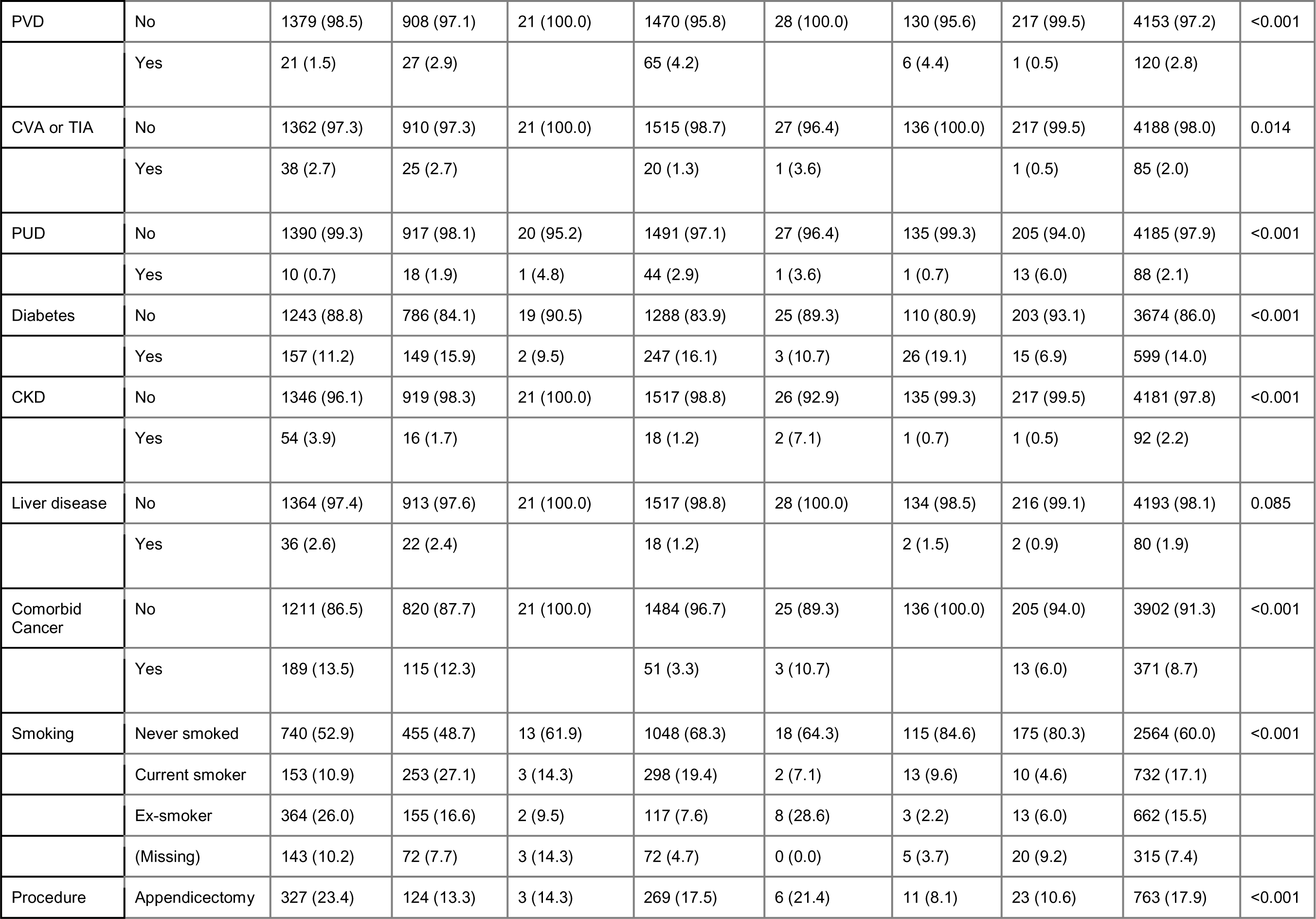
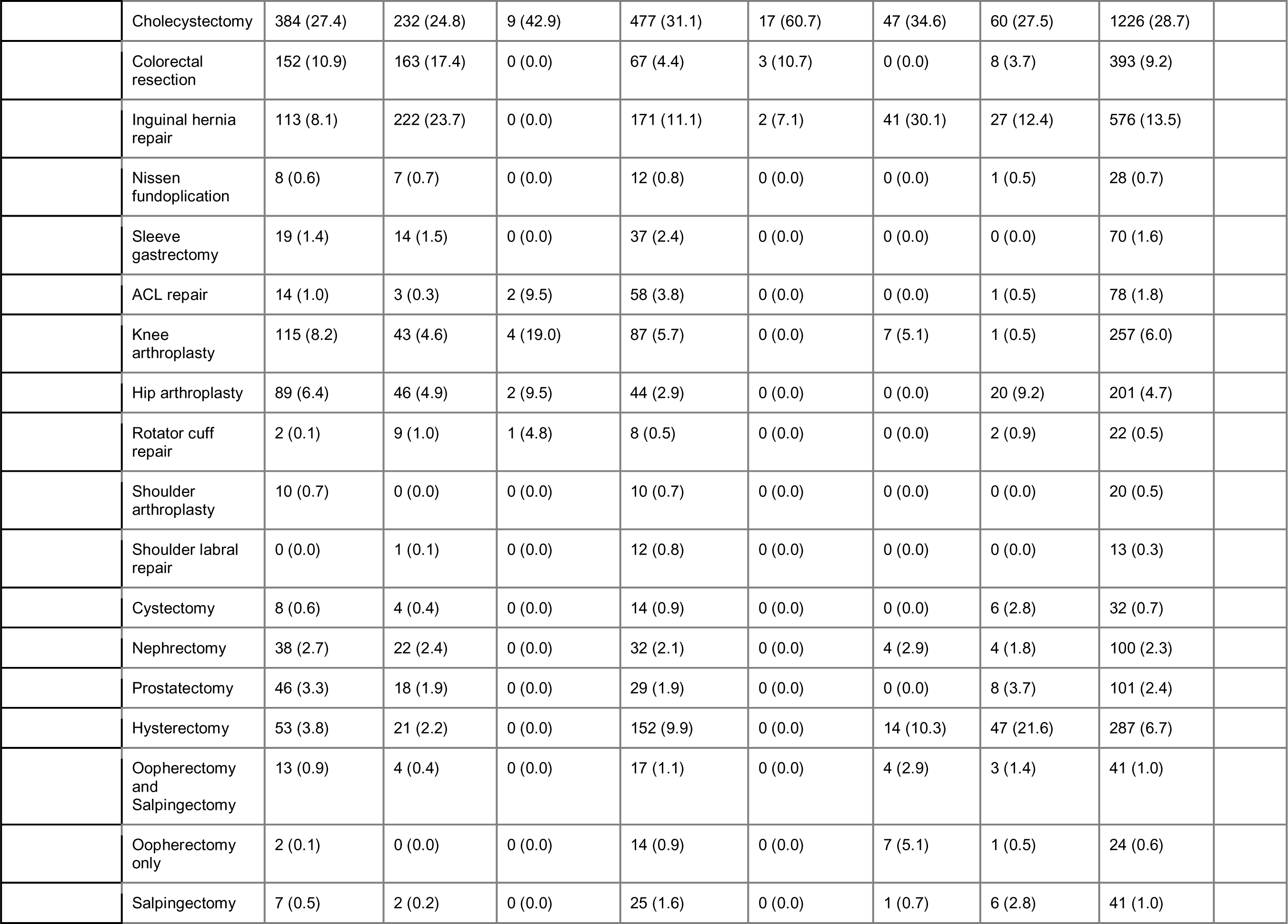
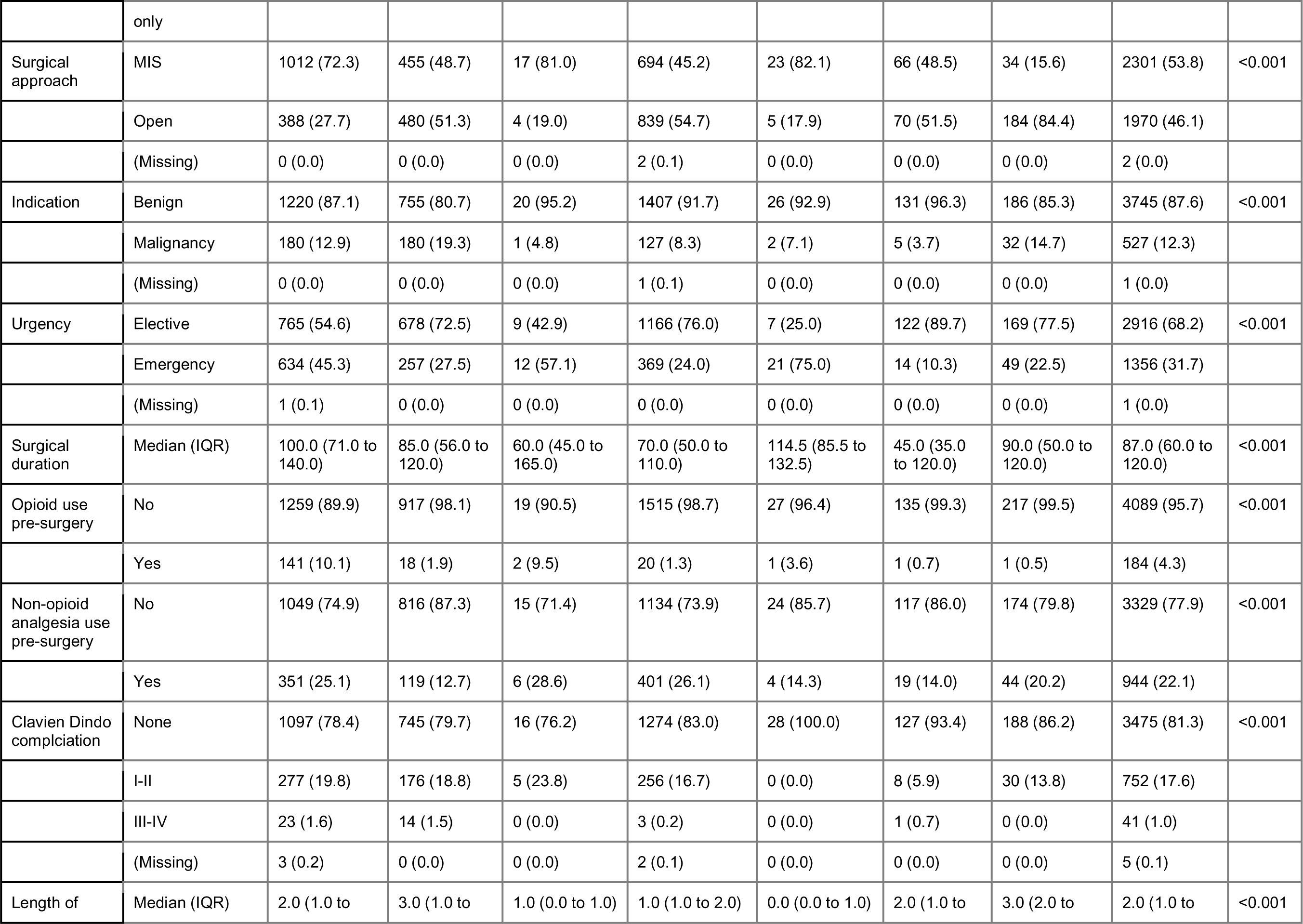
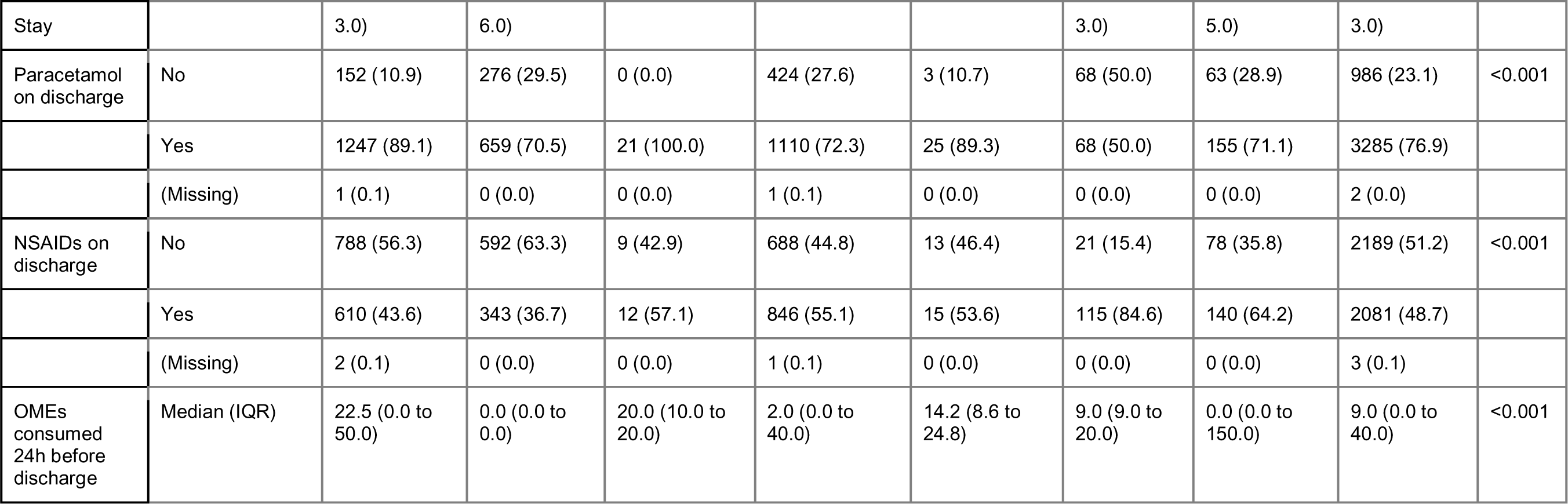
Variations in demographics by region.

**Table S6:**
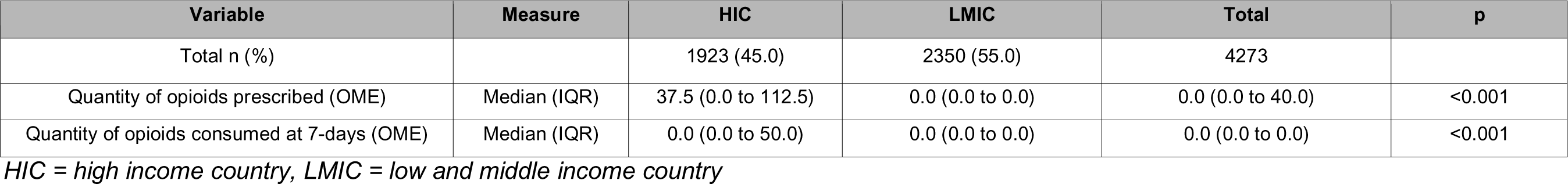
Opioid prescription and consumption quantities by country income group.

**Table S7:** Comparison of those lost-to-follow-up *versus* those included in the final cohort.

| Variable | Label | Lost-to-follow-up | Included Cohort | Total | p |
| --- | --- | --- | --- | --- | --- |
| Total N (%) |  | 957 (18.3) | 4273 (81.7) | 5230 |  |
| Age | Median (IQR) | 53.0 (35.0 to 66.0) | 50.0 (34.0 to 64.0) | 50.0 (34.0 to 65.0) | 0.016 |
| Gender | Female | 521 (54.4) | 2271 (53.1) | 2792 (53.4) | 0.558 |
|  | Male | 436 (45.6) | 1999 (46.8) | 2435 (46.6) |  |
|  | Other |  | 3 (0.1) | 3 (0.1) |  |
| ASA | I | 327 (34.5) | 1763 (41.3) | 2090 (40.1) | <0.001 |
|  | II-III | 605 (63.9) | 2466 (57.8) | 3071 (58.9) |  |
|  | IV - V | 15 (1.6) | 39 (0.9) | 54 (1.0) |  |
| BMI | Normal (18.5 -24.9) | 237 (25.4) | 1162 (27.4) | 1399 (27.0) | <0.001 |
|  | Obese (31-40) | 238 (25.5) | 936 (22.1) | 1174 (22.7) |  |
|  | Overweight (25-30) | 242 (25.9) | 1463 (34.5) | 1705 (32.9) |  |
|  | Severely obese (>40) | 47 (5.0) | 205 (4.8) | 252 (4.9) |  |
|  | Underweight (< 18.5) | 19 (2.0) | 69 (1.6) | 88 (1.7) |  |
|  | Unknown / Not Recorded | 151 (16.2) | 406 (9.6) | 557 (10.8) |  |
| Comorbid cancer | No | 858 (89.7) | 3902 (91.3) | 4760 (91.0) | 0.118 |
|  | Yes | 99 (10.3) | 371 (8.7) | 470 (9.0) |  |
| Smoking | Current smoker | 163 (17.1) | 732 (17.1) | 895 (17.1) | 0.003 |
|  | Ex-smoker < 12 months | 21 (2.2) | 104 (2.4) | 125 (2.4) |  |
|  | Ex-smoker >12 months | 118 (12.4) | 558 (13.1) | 676 (12.9) |  |
|  | Never smoked | 542 (57.0) | 2564 (60.0) | 3106 (59.5) |  |
|  | Unknown status | 107 (11.3) | 315 (7.4) | 422 (8.1) |  |
| Procedure | ACL repair | 10 (1.0) | 78 (1.8) | 88 (1.7) | <0.001 |
|  | Appendicectomy | 160 (16.7) | 763 (17.9) | 923 (17.6) |  |
|  | Cholecystectomy | 291 (30.4) | 1226 (28.7) | 1517 (29.0) |  |
|  | Colorectal resection | 91 (9.5) | 393 (9.2) | 484 (9.3) |  |
|  | Cystectomy | 6 (0.6) | 32 (0.7) | 38 (0.7) |  |
|  | Hip arthroplasty | 62 (6.5) | 201 (4.7) | 263 (5.0) |  |
|  | Hysterectomy | 49 (5.1) | 287 (6.7) | 336 (6.4) |  |
|  | Inguinal hernia repair | 89 (9.3) | 576 (13.5) | 665 (12.7) |  |
|  | Knee arthroplasty | 70 (7.3) | 257 (6.0) | 327 (6.3) |  |
|  | Nephrectomy | 33 (3.4) | 100 (2.3) | 133 (2.5) |  |
|  | Nissen fundoplication | 6 (0.6) | 28 (0.7) | 34 (0.7) |  |
|  | Oophorectomy and Salpingectomy | 6 (0.6) | 41 (1.0) | 47 (0.9) |  |
|  | Oophorectomy only | 4 (0.4) | 24 (0.6) | 28 (0.5) |  |
|  | Prostatectomy | 31 (3.2) | 101 (2.4) | 132 (2.5) |  |
|  | Rotator cuff repair | 4 (0.4) | 22 (0.5) | 26 (0.5) |  |
|  | Salpingectomy only | 5 (0.5) | 41 (1.0) | 46 (0.9) |  |
|  | Shoulder arthroplasty | 19 (2.0) | 20 (0.5) | 39 (0.7) |  |
|  | Shoulder labral repair | 1 (0.1) | 13 (0.3) | 14 (0.3) |  |
|  | Sleeve gastrectomy | 20 (2.1) | 70 (1.6) | 90 (1.7) |  |
| Surgical approach | MIS | 546 (57.5) | 2301 (53.9) | 2847 (54.5) | 0.048 |
|  | Open | 404 (42.5) | 1970 (46.1) | 2374 (45.5) |  |
| Indication | Benign | 821 (86.5) | 3745 (87.7) | 4566 (87.5) | 0.36 |
|  | Malignancy | 128 (13.5) | 527 (12.3) | 655 (12.5) |  |
| Urgency | Elective | 637 (67.1) | 2916 (68.3) | 3553 (68.0) | 0.495 |
|  | Emergency | 313 (32.9) | 1356 (31.7) | 1669 (32.0) |  |
| Specialty | General surgery | 657 (68.7) | 3056 (71.5) | 3713 (71.0) | <0.001 |
|  | Gynaecological surgery | 64 (6.7) | 393 (9.2) | 457 (8.7) |  |
|  | Orthopaedic surgery | 166 (17.3) | 591 (13.8) | 757 (14.5) |  |
|  | Urological surgery | 70 (7.3) | 233 (5.5) | 303 (5.8) |  |
| Discharged with opioid | No | 585 (61.1) | 2962 (69.3) | 3547 (67.8) | <0.001 |
|  | Yes | 372 (38.9) | 1311 (30.7) | 1683 (32.2) |  |

**Figure S1:**
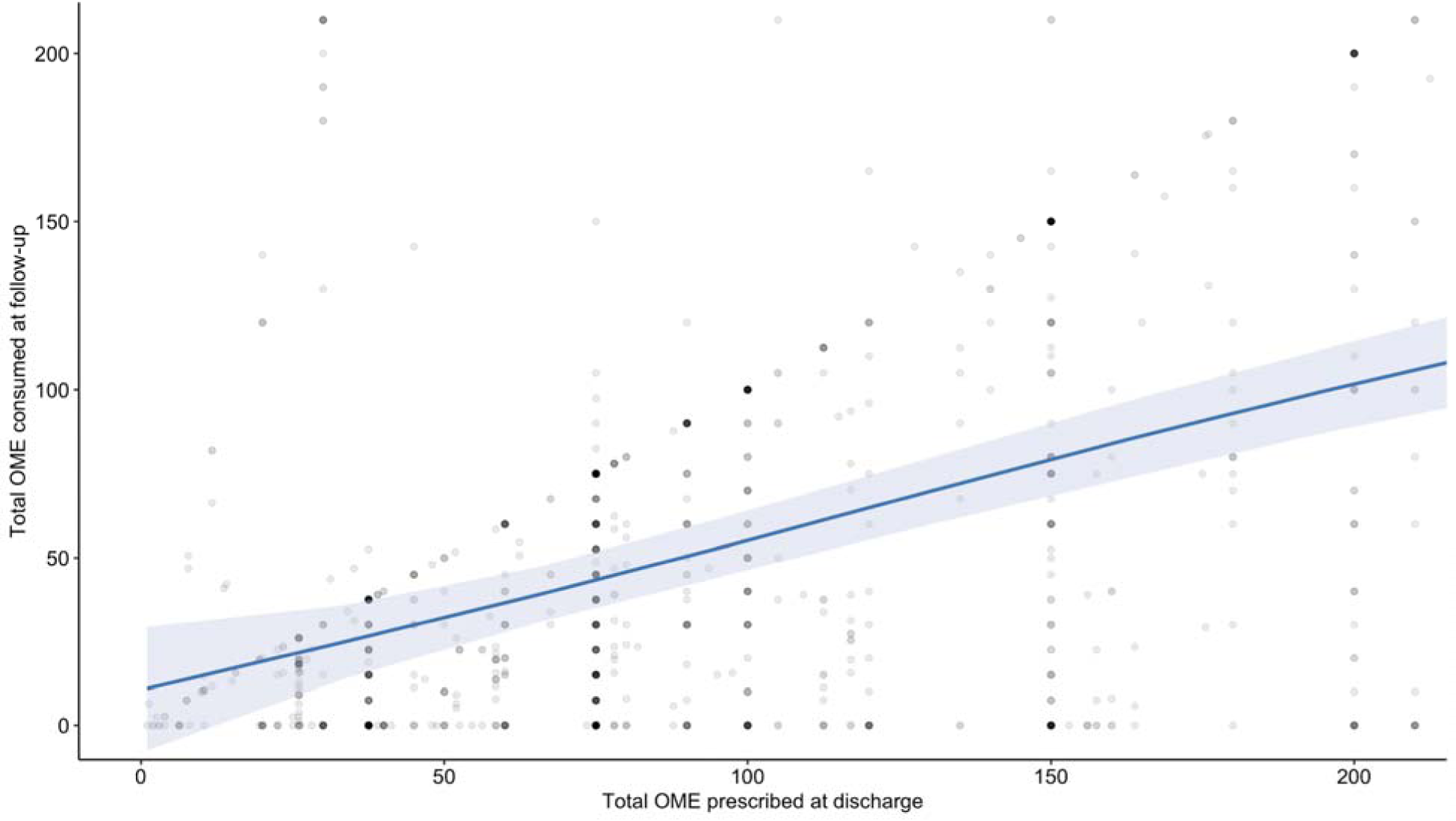
Total amount of oral morphine equivalents (OME) consumed within 7 days and total amount of OMEs prescribed at discharge. Plotted line represents a smoothed conditional mean from a fitted generalised additive model. The shaded area denotes bounds of the 95% CI.

**Figure S2:**
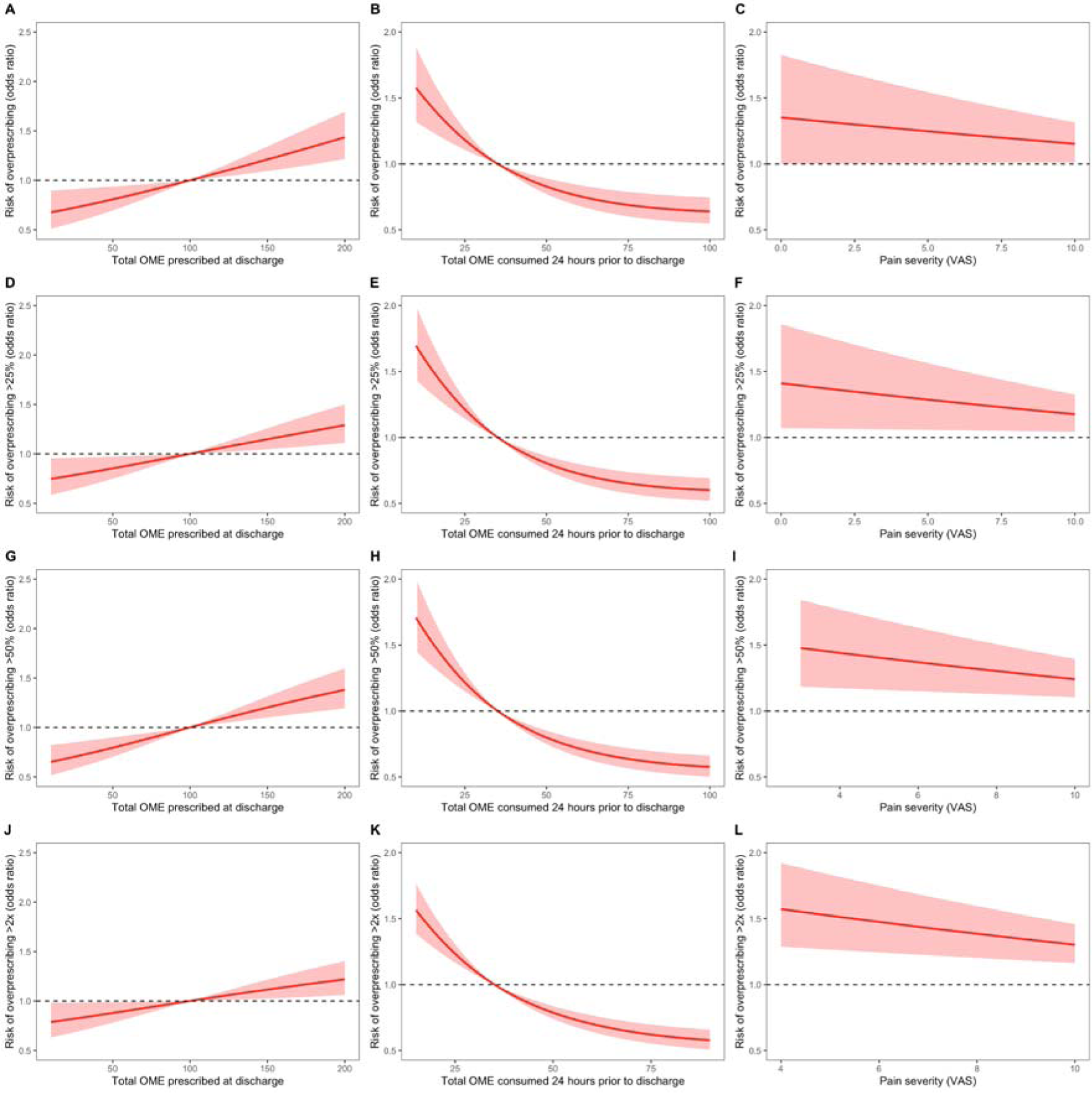
Restricted cubic spline plots with 3 knots for a binary logistic regression model for the risk of over-prescribing opioids (prescribing more than what is consumed at 7-days follow-up). A) Plots risk of overprescription across a spectrum of OMEs prescribed at discharge; B) plots risk of overprescription across a spectrum of OMEs consumed 24 hours prior to discharge; and C) plots risk of overprescription across a spectrum of numeric rating scale pain scores. Sensitivity analyses were performed thresholding differences between prescription and consumption quantities of OMEs (D, E, F: prescriptions 25% more than consumed; G, H, I: prescriptions 50% more than consumed; J, K, L: prescription 100% more than consumed).

### PubMed Citable Authors

#### Writing group

Lorane Gaborit* [Australian National University, Canberra, Australia]; Kaviya Kalyanasundaram [University of Adelaide, Adelaide, Australia]; Jennifer Vu [Sydney University, Sydney, Australia]; Aya Basam [Monash University, Melbourne, Australia]; Muhammed Elhadi [University of Tripoli, Tripoli, Libya]; Deborah Wright [University of Otago, Otago, New Zealand]; Jennifer Martin, Melissa Park, Peter Pockney [University of Newcastle, Newcastle, Australia]; Maria Ntalouka [Larissa University Hospital, Larisa, Greece]; Noora Abubaker [Ibn-Sina Hospital, Khartoum, Sudan]; Muhammed Elhadi [University of Tripoli, Tripoli, Libya]; Umar Saeed [International Center of Medical Sciences Research, Islamabad, Pakistan]; Eman Abdulwahed [University of Tripoli, Tripoli, Libya]; Mohamed Alsori, Ghaliya Mohamed H Alrifae [Tripoli Medical Center/ Tripoli University Hospital, Tripoli, Libya]; Michael Farrell [Lehigh Valley Health Network, Pennsylvania, United States of America]; Gordon Liu, Nicholas Smith, William Xu, Chris Varghese** [University of Auckland, Auckland, New Zealand]

*First author

**Senior author (overall guarantor)

#### Statistical analysis

Chris Varghese** [University of Auckland, Auckland, New Zealand], Ewen Harrison [University of Edinburgh, Edinburgh, United Kingdom]

#### OPERAS Steering committee

Aya Basam, Sarah Goh, Jiting Li, Jainil Shah, Abdullah Waraich [Monash University, Melbourne, Australia]; Lorane Gaborit, Upasana Pathak [Australian National University, Canberra, Australia]; Amie Hilder [Deakin University, Melbourne, Australia]; Muhammed Elhadi [University of Tripoli, Tripoli, Libya]; Aiden Jabur [Griffith University, Gold Coast, Australia]; Kaviya Kalyanasundaram [University of Adelaide, Adelaide, Australia]; Christina Ohis [Western Sydney University, Sydney, Australia]; Chui Foong [Kelly] Ong [Melbourne Training Circuit, Melbourne, Australia]; Melissa Park, Venesa Siribaddana [University of Newcastle, Newcastle, Australia]; Kyle Raubenheimer [Perth Metro Training Circuit, Perth, Australia]; Jennifer Vu [Sydney University, Sydney, Australia]; Cameron Wells, Gordon Liu, Liam Ferguson, William Xu, Chris Varghese [University of Auckland, Auckland, New Zealand]

#### Scientific advisory group

Peter Pockney, Kristy Atherton, Amanda Dawson, Jennifer Martin [University of Newcastle, Newcastle, Australia]; Arnab Banerjee [Australian National University, Canberra, Australia]; Nagendra Dudi-Venkata [Royal Australasian College of Surgeons, Adelaide, Australia]; Nicholas Lightfoot [University of Auckland, Auckland, New Zealand]; Isabella Ludbrook [Hunter New England Network, Newcastle, Australia]; Luke Peters [Royal Australasian College of Surgeons, Sydney, Australia]; Rachel Sara [Counties Manukau Health, Manukau City, New Zealand]; David Watson [Flinders University, Adelaide, Australia]; Deborah Wright [University of Otago, Otago, New Zealand]

#### OPERAS National Leads

Ademola Adeyeye [Afe Babalola University Multisystem Hospital, Ado-Ekiti, Nigeria]; Luis Adrian Alvarez-Lozada [Autonomous University of Nuevo León, Monterrey, Mexico]; Semra Demirli Atici [University of Health Sciences Tepecik Training and Research Hospital, Izmir, Turkey]; Milos Buhavac [Texas Tech University Health Sciences Center, Lubbock, United States of America]; Giacomo Calini [University Hospital of Udine, Udine, Italy]; Muhammed Elhadi [University of Tripoli, Tripoli, Libya]; Orestis Ioannidis [George Papanikolaou Hospital, Thessaloniki, Greece]; Mustafa Deniz Tepe [Karadeniz Technical University, Trabzon, Turkey]; Upanmanyu Nath [Nilratan Sircar Medical College and Hospital, Kolkata, India]; Ahmad Uzair [King Edward Medical University Hospital, Lahore, Pakistan]; Wah Yang [The First Affiliated Hospital of Jinan University, Guangzhou, China]; Faseeh Zaidi, Surya Singh [University of Auckland, Auckland, New Zealand]; Bahiyah Abdullah [Hospital Universiti Teknologi MARA, Malaysia (HUiTM)], Diana Sofia Garces Palacios* [Hospital Susana Lopez De Velencia], Ahmed Ragab [Alexandria University, Alexandria, Egypt]

#### OPERAS Australian State Leads

Kyle Raubenheimer [Royal Perth Hospital, Perth, Australia]; Davina Daudu [University of Western Australia, Perth, Australia]; Sarah Goh, Simran Vinod Benyani, Nandini Karthikeyan [Monash University, Melbourne, Australia]; Laure Taher Mansour [University of Adelaide, Adelaide, Australia]; Warren Seow [University of Adelaide, Adelaide, Australia]; Zoya Tasi [University of Tasmania, Hobart, Australia]; Aiden Jabur [Griffith University, Gold Coast, Australia]; Upasana Pathak [Australian National University, Canberra, Australia]; Melissa Park [University of Newcastle, Newcastle, Australia]

#### Algeria

Dhia Errahmane Abdelmelek*, Ikram Fatima Zohra Boussahel, Oumelaz Kaabache, Naoual Lemdaoui, Oualid Nebbar [Center Anti Cancer, Sétif]; Mounira Rais*, Meriem Abdoun*, Aya Tinhinane Kouicem, Souad Bouaoud, Kamel Bouchenak, Hind Saada, Amel Ouyahia, Wassila Messai [CHU Saadna Abdennour Hospital, Sétif]

#### Australia

Zhi Shyuan Choong*, Clarissa Ting, Michelle Larkin, Pei Jun Fong, Isabel Soh, Alyssia De Grandi, Hareem Iftikhar, Akansha Sinha, Dhruv Kapoor, Tara Chlebicka [Albury Wodonga Health]; David Singer*, Kim Goddard, Lisa Matthews [Armadale Health Service, Mount Nasura]; Rosalina Lin*, Jessica Chambers, Juliet Chan, Brooke Macnab, John Barker, Morgan Mckenzie, Neil Ferguson [Armidale Rural Referral Hospital, Armidale]; Ghanisht Juwaheer*, Vijayaragavan Muralidharan, Sonia Gill, Nakjun Sung, Rohan Patel, Chris Walters, Kevin Nguyen, David Liu, Carlos Cabalag, Jennifer Lee, San-Hui Anita Leow, Suat Li Ng, Hamza Ashraf, Fraizer Mulder, Jonathan Loo, David Proud, Samantha Wong, Yida Zhou, Qi Rui Soh, David Chye, Sean Stevens, Patrick Tang, Stephen Kritharides, Jason Dong, Oscar Morice, Dora Huang, Andrew Hardidge, Mishka Amarasekara, Aleah Kink, Damien Bolton [Austin Hospital, Melbourne]; Alisha Rawal*, Jasraaj Singh*, Matthew Heard*, Yusuf Hassan*, Ahmed Naqeeb, Andrew Cobden, Duron Prinsloo, Dwain Quadros, Emma Gunn, Ha Jin Kim, Jennifer Ekwebelam, James Shanahan, Mustafa Alkazali, Mariyah Hoosenally, Naveen Nara, Peter Nguyen, Sally Barker [Ballarat Base Hospital, Ballarat]; Amie Hilder*, Ally Hui, Antara Karmakar, Bill Wang, Janindu Goonawardena, King Tung Cheung, Nicholas Chan, Ragul Natarajan, Richard Cade, Rong Jin, Shomik Sengupta, Ruth Snider [Box Hill Hospital, Melbourne]; Harsha Morisetty*, Lewis Weeda, Phoebe Sun, Lalitya Chilaka, Jacinta Cover [Bunbury Hospital]; Aashrinee De Silva Abeweera Gunasekara*, Rahavi Senthilrajan, Anas Alwahaib, Alexandra Limmer, Bushra Zamanbandhon [Campbelltown Hospital]; Kumail Jaffry [Casey Hospital, Melbourne]; Yijia Shen*, Alan Chua, Saifulla Syed [Central Gippsland Health]; Sushanth Saha*, John Glynatsis*, Lori Aitchison, Bernard Lagana, Mason Crossman, David Watson, Abby Dawson, Bryan Fong, Ella Harrison, Eleanor Horsburgh, John Glynatsis, Michael Khoo, Kritika Mishra, Lewis Hewton, Alex Mesecke, Hien Tu, Than Tun, Jason Wong [Flinders Medical Centre, Adelaide]; Elynn Ong*, Tara-Nyssa Law*, Ashlee Landy, Alyssa Leano, Andrea Li, Akshay Soni, Benjamin Dowdle, Charles Pilgrim, Dewmi Abeysirigunawardana, Deepak Rajan Jeyarajan, Diya Patel, Kyle Mckinnon, Madeline Gould, Paul Gilmore, Ruxi Geng, Rachael Loughnan, Sarahjane Norton-Smith, Solomon Nyame, Sarah Tan, Si Woo Yoon, Yantong Wang, Yichi Zhang, Zixuan Wang [Frankston Hospital]; Hans Mare*, Indrajith Withanage [Geraldton Regional Hospital]; Mitali Khattar*, Alexandra Toft, Goutham Sivasuthan, Hailin Zhao, Jordan Addley, Lucinda O’brien, Muhammad Raza, Randipsingh Bindra, Sonakshi Sharma [Gold Coast University Hospital, Southport]; Charlotte Cornwell*, Aditya Patil, Aiden Cheung, Ashleigh Lown, Amanda Dawson, Aneel Blassey, Benjamin Ochigbo, Felicity Cheng, Aleeza Fatima, Edward Zhang, Henry Kocatekin, Charles Roth, Dani Brewster, Kelvin Kwok, Paul Chen, Sharon Laura, Dominic Tynan, Edward Latif, Elizabeth Lun, Elodie Honore, Felix Ziergiebel, Jessica Blake, Karan Chandiok, Katie Bird, Lynette Ngothanh, Melissa Lee, Mariam El-Masry, Peter Hamer, Ramanathan Rm Palaniappan, Richard Mcgee, Sarah Huang, Shane Zhang, Shubhang Hariharan, Yannick De Silva, Celeste Lee, Penelope Fotheringham, Ian Incoll, Timothy Cordingley, Felicity Cheng, Matthew Brown, Leannedra Kang, Rivindu Wijayaratne, Parisse Moore, Gemma Qian, Yara Elgindy [Gosford Hospital, Gosford]; Emma Carnuccio*, Hamish Rae, Mena Shehata [Goulburn Base Hospital]; Mingchun Liu*, Brodee Lockwood, John Van Bockxmeer [Hedland Health Campus]; Ali Alsoudani*, Daniel Swan, Justin Hsieh [Ipswich Hospital]; Francesca Orchard-Hall*, Kai Yun Jodene Tay*, Raagini Mehra*, Alpha Gebeh, Ashley Bailey, Georgia Brown, Ashley Colaco, Hemashree Gopal, Jessica Boyley, Varun Changati, Joseph Fletcher, Tanishq Khandelwal, Colin House, Chris O’neil, Emily Jaarsma, Victor Ly, Zsolt Balogh, Amanda Shui, Vinogi Sathasivam, Hannah Legge-Wilkinson [John Hunter Hospital, Newcastle]; King Ho Wong*, Andrew Chen, Anthony Tran, Peter Rehfisch, Grace Wang, Jonathan Nguyen, Joshua Peker, Kayla Gallert, Mia Komesaroff, Manideep Namburi [Latrobe Regional Hospital]; Elisabeth Goldfinch*, Ropafadzo Muchabaiwa*, Aishwarya Jangam, Isobel Taylor, Iulian Nusem, Jin Hyuk (David) Park, Justin Gundara, Rachael Heigan, Tam Tran, Thomas Mackay, Yasmine Butterworth, Tomas Sadauskas, Melody Tung, Hasthika Ellepola [Logan Hospital, Meadowbrook]; Christine Gan*, Hakim Fong*, Ankita Das, Leshya Naicker, Samantha Hauptman, Aditi Kamath, Anthea Yew, Anupam Parange, Katie Kim, Sahil Kharwadkar, Tharushi Gamage [Lyell Mcewin Hospital]; Lucille Vance*, Alexandra Seldon, Moheb Ghaly [Manning Base Hospital]; Victoria Phan [Maroondah Hospital, Melbourne]; Karanjeet Chauhan*, Ahmad Bassam, Beverley Vollenhoven, Kumail Jaffry, Kajal Mandhan, Mithra Sritharan, Mahesh Sakthivel, Natalie Evans, Samuel Robinson, Seiyon Sivakumar [Monash Medical Centre, Clayton]; Liberty Marrison*, David Jollow, Krishma Joshi, Steve Tao, Pallavi Shrestha, Sai Keerthana Nukala [Northern Beaches Hospital]; Russell Hodgson*, Anna Crotty, Adriana Esho, Alasdair Harris, Amy Surkitt, Laura Bland, Blake Mcleod, Chonghao Yin, Cambo Keng, Emily Greenwood, Grace Yuan, Emma Haege, Hongyi Wu, Haotian Xiao, Isabella Pozzi, Jeff Fu, Jessica Stott Ross, Juliette Gentle, Kathy Gan, Kelvin Chang, Kexin Sun, Madhavi Singh, Maria Xie, Nicholas Mccabe, Mark Slavec, Nick Clarnette, Behzad Niknami, Peishan Zou, Sean Flintoft, Shenuka Jayatilleke, Rumnea Sok, Suqi Tan, Sanya Wadhwa, Will Swansson [Northern Hospital, Melbourne]; Daniel Abulafia*, Jian Blundell*, Amie Sweetapple, Caitlin Del Solar, Cameron Martin, David Bell, Isuru Fernando, Jared Chang, Katie Vanzuylekom, Katie Van Zuylekom, Kate Van Zuylekom, Katie Hobbs, Richard Liang [Orange Base Hospital]; Aiden Jabur*, Jazmina Tarmidi, Mahmoud Ugool, Nicholas Beatson, Sarah Bowman, Sophie Moin [Queen Elizabeth Ii Jubilee Hospital, Coopers Plains]; Wen Po Jonathan Tan*, Seevakan Chidambaram*, Siang Wei Gan, Pengnan Wang, Leshya Naicker, Katie Kim, Nicole Qiwen Wang, Yi Xin Kwan, Chinmai Patil, Divyanshu Joshi, Aditi Kamath, Aishath Hanan, Arfaan Sheriff, Jaime Duffield, Leshya Naiker, Peter Smitham, Eu Ling Neo, Matthew Chua, Shalvin Prasad, Armitesh Nagaratnam, Tarik Sammour, Yuxin Lin, Christine Lee, Eve Hopping, Muskan Jangra, Ankita Das, Ken Lin, Zachary Bunjo [Royal Adelaide Hospital, Adelaide]; Kyle Raubenheimer*, Mohamed Haseef Mohamed Yunos, Kar Long Yeung, Rachel Phu, Aisling Betts, Benjamin Just, Sahil Gera, Hilary Leeson, Jodie Jamieson, Katie Wang, Emily Luu, Michael Innes [Royal Perth Hospital]; Jennifer Vu*, Jonathan Hong, Stephen Dzator, Aki Flame, Vincent Jiang, Jianing Kwok, Aaron Lawrence, Kate Meads, Liam Pearce, Pavatharane Sarangadasa, Haylee Shaw, Victor Yu, [Royal Prince Alfred Hospital]; Elizabeth Crostella*, James Wong, Sriya Bobba, Maddison Muller, Yin Chi Hebe Hau, Thomas Wilson, Aleksandra Markovic, Jemma Green, Clara Forbes, Emalee Burrows, Lachlan Hou, Clare O’sullivan, Jonathon Foo [Sir Charles Gairdner Hospital, Perth]; Hannah Greig*, A-J Collins, Callum Chandler, Emily Heaney, Hannah Gross, Monica Morgan, Rebecca Loder, Krishnankutty Rajesh [New South Wales Local Health District Site Bega Hospital]; Shravankrishna Ananthapadmanabhan*, Akeedh Razmi, Crystal Vong, Prasanna Pothukuchi, Mary Theophilus, Roshni Sriranjan, Sharon Kaur, Marcelo Kanczuk [St John Of God Midland Public And Private Hospital, Perth]; Julia De Groot*, Angela Corrigan, Damon Li, Danniel Badri, Dominico Ciranni, Elangovan Thaya Needi, Matthew Clanfield, Nicolas Copertino, William Rumble [Sunshine Coast University Hospital]; Maria Kristina Vanguardia*, Chen Lew*, Rami Dennaoui*, Jainil Shah*, Joseph Kong, Imogen Koh, Raymond Zeng, Kristian Baziotis-Kalfas, Hannah Denby, Andy Li, Will Tran, Abhinav Singh, Olivia Lin, Michelle Chau, Olivia Donaldson, Christina (Seojung) Min, Shirahn Ballah, Sonia Ching Ting Tsui, Nathania Yong, Lucy Standish, Sarah Tan, AsukaFujihara, Lily Davies, Ramin Odisho, Anjana Ravi, Josh Collins, Pooja Chandra, Rana Abdelmeguid, GopalSingh, Xireaili Feierdaiweisi, Dharani Seneviratne, Shambhavi Srivastava, Michelle Yao, Cherilyn Teng, Nebula Chowdhury, Sasini Vidanagama, Charles Lin, Tharushi Sampatha-Waduge, Erica Wang, Chatnapa Yodkitydomying, Imogen Koh, Julia Silverii, AaronLam, Raymond Zeng, Krisha Solanki, Angus Franks, Liam Edwards, Ridvan Atilhan, Rohan Nandurkar, Oliver Wells, Kristina Vanguardia, Dennis King, Elton Edwards, Liam Edwards, Quang Tran, Michelle Chau, Seojung Min [The Alfred Hospital, Melbourne]; Abdul Rauf*, Yangzirui Fu*, Hodo Haximolla, Mengge Shang, Sharrada Segaran, Shelley Wang, Gananadha Sivakumar [The Canberra Hospital]; Jaspreet Kaur Sandhu*, Neel Mishra, Samantha Hauptman, Alyssa Chua, Danielle Chene, Guy Maddern, Henry Shaw, Qiwen Wang [The Queen Elizabeth Hospital, Adelaide]; Siyuan Pang*, Christine Lu, James Fung, Kathryn Cyr, Karen Lu, Ming Zhou How, Nelson Hu, Paul Anderson, Philip Jakanovski [The Royal Melbourne Hospital, Melbourne]; Arkan Youssef*, Howard Tang*, Rory Keenan*, Alex Chan, Mitch Canny, Farah Tahir, James Egerton, Justin Yeung, Justin Chan, Lea Tiffany, Michael Bei, Mariolyn Raj, Peter Williams, Sakshar Nagpal, Tim Outhred, Russel Krawitz [Western Health, Melbourne]; Khadijah Younus*, Mary Giurgius*, Rosemary Kirk, Amanda Gonzalez Pegorer, Pattarapan Tang-Ieam, Jack Ward, Asanka Wijetunga, Caitlin Zhang, Chris Nahm, Christine Wang, Damian Golja, Gregory Jenkins, Helena Qian, Jason Luong, Kim Nguyen, Sean Suttor, Sherman Lai, Vanessa Ma, Yan Chen [Westmead Hospital, Westmead]; Hoi Hang Yu*, Amos Lee, Antonio Barbaro, Cameron Mcguinness, Guy Maddern, Stevie Young [Whyalla Hospital & Health Services, Whyalla]; Ye Fang Lim*, Georgina Trotta, Phoebe Chao, George Ding, Carol Fang, Andi Lu, Prabhath Wagaarachchi [Women’s And Children’s Hospital]; Charlotte Cornwell*, Amy Gojnich, Peter Stewart, Isabella Dong, Kenneth Wong, Luca Burruso, Lucinda Hogan, Nathan Mcorist, Ramnik Singh, Ragavi Jeyamohan, Zhen Hou, William Lai, Emily Taylor [Wyong Public Hospital, Wyong]

#### Colombia

Diana Sofia Garces Palacios*, Maria Alejandra Nanez Pantoja, Daniel Mauricio Bolanos Nanez, Gilmer Omar Perez Hernandez, Lia Jasmin Jimenez Ramirez [Hospital Susana Lopez De Velencia]

#### Egypt

Mohamed Mohamed*, Ahmed Kamal El-Taher, Ahmed Elewa, Mahmoud Ayman Soliman, Menna Diab, Radwa Ali [Al Tayseer Hospital, Zagazig]; Ahmed Ahmed*, Adham Galal, Ahmed Elkhodary, Ali Alaa, Arwa Faisal, Asmaa Badawy, Donia Eldomiaty, Mohamed Al Sayed, Esraa Rasslan, Mohamed Ramadan, Gamal Elsayed Fares, Hashem Altabbaa, Humam Emad, Muneera Alboridy, Mahmoud Mongy, Osama Albarhomy, Osama Selim, Rawan Rafaei, Raneem Atta, Ahmad Altaweel, Yara Sherif, Youssef Elghoul, Yousef Tarek [Alexandria Main University Hospital, Alexandria]; Dina Atef*, Ahmed Mahmoud*, Mahmoud Saad*, Mohamed Ragab, Aya Hussien, Mostafa Abdelbaky, Ismail Muhammad, Afnan Morad, Ahmed Ali, Ahmed Hussien, Ahmed Shipa, Ahmed Aboulfotouh, Ahmed Mohamed Hashem, Ahmed Morsi, Alshymaa Ebrahim, Ahmed Mohamed Sayed, Amira Abdelrahman, Aml Ali, Samah Abdelnaeam, Asmaa Emam, Aya Shaban, Fady Barsoum, Esraa Mostafa, Doaa Abdelbaset, Dina Othman, Safaa Othman, Nour Salah Khairallah, Salma Morsi, Armia Azer, Enas Abdelbaset Abdelsamed, Islam Ibrahim, Esraa Abdelbaset, Esraa Hamoda, Fatma Monib, Fatma Harb, Hager Maher, Haitham Mohammed, Kerollos Henes, Kerollos Shamshoon, Mahmoud Hassanein, Magdy Mahdy, Mahmoud Khalil, Manal Ali, Mansour Khalifa, Marwa Amary, Merna Ezz Suliman, Mohammed Saif Al Nasr, Michael Elia, Michael Adly, Mo’men Roshdy, Mohammed Al-Quossi, Mohammed Fargaly, Mona Saber, Mostafa Abbas, Ola Haroon, Omima Khalil, Omnia Talaat, Rahma Elnagar, Randa Soliman, Reham Aboelela, Salem Salah, Samia Abdelgawad, Tarek Hussien, George Sobhy, Yasmeen Sayed, Yousra Othman [Assiut University Hospital, Assiut]; Reham Silem*, Ali Dawood, Tarek Hemaida, Reem Ahmed [Aswan University Hospital, Aswan]; Ebrahim Salem, Osama Fathy Ali Ali Rashed, Mohamed Halawa [El Tadamon Specialised Hospital, Portsaid]; Hossam Elfeki*, Abdelrahman Mosaad, Abdelrahman Shaaban, Hebatalla Abdelsalam, Ahmed Sakr, Aly Sanad, Amr Elsawy, Bassant Maged Maged, Dana Hegazy, Mohamed Abdelmaksoud, Mahmoud Laymon, Mohamed Taman, Esraa R Moawad, Hadeer Aboelfarh, Karim Elkenawi, Manar Osama, Mirna Sadek, Mohamed Abdelaziz Elghazy, Mohammed Attiah, Mohamed Nader, Mostafa Shalaby, Omar Attiya, Osama Samir Gaarour, Ahmed Zaghloul [Mansoura University Hospital, Mansoura]; Pola Mikhail*, Karim Badr, Hatem Soltan, Mohamed Donia, Mohammed Gaafar [Menofia University Hospital, Menofia]; Khaled Abdelwahab*, Abdelaziz Sallam, Ahmed Eid, Mohamed Yousri, Omar Hamdy [Oncology Center Mansoura University, Mansoura]; Aiman Al-Touny*, Abdelrhman Alshawadfy, Ahmed Hamdy, Ahmed Ellilly, Ahmed Mahdy, Ahmed El-Sakka, Hamdy Hendawy, Asmaa Salah, Bassma Raslan, Eman Teema, Eslam Albayadi, Esraa Nasser, Hanaa Mohamed, Mohamed Mahmoud, Mostafa Elsaied, Omima Taha, Shaimaa Dahshan, Shimaa Al-Touny, Ahmed Karrar, Ahmed Khairy [Suez Canal University Hospital, Ismailia]; Alaa Mohamed Ads*, Rabiaa Alomar, Issa AbuShawareb, Abdallah Saeed, Abdelhafeez Mashaal, Adel Mohamed Ads, Sohila Ghanem, Ahmed Elghamry, Eman Ayman Nada, Youssef Ali Noureldin, Mohamed Fayez Fouda, Nourhan Shaheen, Shereen Allam, Ibrahim Mazrou, Ali Fahmy Shehab, Wesam Kussaili [Tanta University Hospital, Tanta]

#### Greece

Dimitrios Korkolis*, Evangelos Fradelos, Aikaterini Sarafi [Agios Savvas Anticancer Hospital Of Athens]; Nikolaos Machairas*, Konstantinos S. Giannakopoulos, Fotios Stavratis, Georgios Korovesis, Gerasimos Tsourouflis, Myrto D. Keramida, Nikolaos Kydonakis, Stylianos Kykalos, Athanasios Syllaios, Panagiotis Dorovinis, Dimitrios Schizas [General Hospital Of Athens “Laiko”]; Orestis Ioannidis*, Anastasia Malliora, Elissavet Anestiadou, Konstantinos Zapsalis, Fotios Kontidis, Lydia Loutzidou, Nikolaos Ouzounidis, Stefanos Bitsianis, Savvas Symeonidis, Smaragda Skalidou, Orestis Ioannidis, Olga Maria Valaroutsou [General Hospital Of Thessaloniki “George Papanikolaou”]; Themistoklis Dagklis*, Alexandra Arvanitaki, Apostolos Mamopoulos, Apostolos Athanasiadis, Stergios Kopatsaris, Ioannis Kalogiannidis, Ioannis Tsakiridis, Georgios Kapetanios, Evangelos Papanikolaou, Nikolaos Tsakiridis, Fotios Zachomitros [Hippokratio General Hospital Of Thessaloniki]; Andreas Larentzakis*, Argyrios Gyftopoulos, Konstantinos Albanopoulos, Apostolos Champipis, Christos Yiannakopoulos, Gavriella Zoi Vrakopoulou, Konstantinos Saliaris, Konstantinos Lathouras, Spyridon Skoufias, Georgia Doulami [Iaso]; Metaxia Bareka*, Eleni Arnaoutoglou, Fragkiskos Angelis, Fragkiskos Angeslis, Michael Hantes, Maria Ntalouka [Larissa University Hospital]

#### Iraq

Maytham A. Al-Juaifari*, Mohammed Alwash, Rasool Maala, Yasir Adnan Zwain, Sara Ahmed Saleh, Mohammed Khorsheed [Al-Najaf Al-Ashraf Teaching Hospital, Najaf]

#### Italy

Antonio Pesce, Carlo V. Feo*, Massimiliano Bernabei*, Francesca Petrarulo, Nicolò Fabbri, Raffaele Labriola, Silvia Jasmine Barbara [Azienda Unità Sanitaria Locale di Ferrara-University of Ferrara, Ferrara]; Simone Bosi*, Angela Romano, Anna Canavese, Caterina Catalioto, Claudio Isopi, Cristina Larotonda, Gerti Dajti, Matteo Rottoli, Iris Shari Russo, Stefano Cardelli [Irccs Azienda Ospedaliero, Bologna]; Francesco Castagnini*, Francesco Traina, Giulia Guizzardi, Giulia Giuzzardi, Mara Gorgone, Marco Maestri [IRCCS Istituto Ortopedico Rizzoli, Bologna]; Pasquale Cianci*, Ivana Conversano, Enrico Restini, Domenico Gattulli, Giorgia Grillea, Marco Varesano [Lorenzo Bonomo, Andria]; Giacomo Calini*, Adelaide Andriani, Davide Gattesco, Giovanni Terrosu, Mattia Zambon, Pietro Matucci Cerinic, Luisa Moretti, Davide Muschitiello, Samantha Polo, Vittorio Bresadola [University Hospital Of Udine, Udine]

#### Jordan

Salah Abu Wardeh**\***, Mahmoud Al-Baw*, Saif Alhaleeq*, Subhi Al-Issawi*, Abdalqader Al Smadi, Esmat Alsaify, Farah Banihani, Noor Massadeh, Nada Massadeh, Dima Al-issawi, Basel Elyan, Qotadah Al-Shami, Yazan Alomari [Al Basheer Hospital, Amman]; Abed Alazeez Alkhatib*, Bader Alzghoul, Ahmad Saleh, Jamal Yaghmour, Mahmoud Shahin, Mohammed Maali [Al Istiklal Hospital, Amman]; Dawood Alatefi*, Heba Al-Smirat, Abdulhakim Hezam, Nassar Alathameen [Alkarak Governmental Hospital, Alkarak]; Amr Al Hammoud*,Abdulrahim Al Kaddah*, Salem Ayasrah, Hamza Abuuqteish, Tesneem Al-Mwajeh, Reena Makableh, Saad Bataineh, Amin Shabaneh, Wesam Alnatsheh, Marwan Aldeges, Huda Hamad, Sireen Shehahda, Dima Khassawneh, Osama Alzyoud, Risan Alrosan, Hasan Awad, Tariq Khaldoon, Rabab Shannaq, Mohammad Al hamoud, Bader Abo fadalah, Mo’ath Al-Hazaimeh, Wail Khraise [King Abdullah University Hospital, Irbid]; Lara Alnajjar*, Majjd Alnajjar*, Sohaib Al-Omary*, Adnan Ababneh, Alaa Albashaireh, Mohammad Khadrawi, Mohammad Aljamal*, Tayseer Athamneh, Ro-a Muqbel, Maryam Al-jammal, Ahmad Masarrat, Alia Al-zawaydeh, Ibrahim Taha, Taima’ Qattawi, Rayyan Smadi, Ayah Alhaleem, Mosab Alboon, Omar Hazaymeh, Leen Karasneh, Safa’ Al-Haek [Princess Basma Teaching Hospital, Irbid]

#### Libya

Marin Almahroush*, Tamam Alfrijat, Aya Elporgay, Hadeel Shanag, Hamza Agilla, Hind Alameen, Marya Bensalem, Mawadda Altair, Malak Ghemmied, Rehab Alarabi, Sara Alhudhairy [Abu Saleem Trauma Hospital, Tripoli]; Rima Gweder*, Amal Alzarroug, Ebtihal Alabed, Fadwa Elreaid, Omar A Elkharaz, Fatma Fathi Elreaid, Safa Sasi Albatni [Alkhadra Hospital, Tripoli]; Haitham Elmehdawi*, Milad Gahwagi*, Ayman Mohamed, Tariq Alfrjani, Khaled Khafifi, Ayat Rasheed, Ayoub Akwaisah, Hassan Bushaala, Mustafa Elfadli, Mohamed Moftah, Salima Algabbasi, Salma Esaiti, Sara Elfallah, Abtisam Alharam, Fatima Alariby, Mohamed Isweesi, Tarik Ahmed Eldarat, Ayman Arhuma Dabas [Benghazi Medical Center, Benghazi]; Akram Alkaseek*, Ahmed Mohammed Abodina, Aya Alqaarh, Hibah Bileid Bakeer, Hoda Salem Alhaddad, Husein Aboudlal, Sawsan Alsaih [Gharyan Central Hospital, Gharyan]; Noora Abubaker, Najwa Abdelrahim*, Ali Alzarga, Basma Omar, Farah Faris, Qamrah Alhadad [Ibn Sina Teaching Hospital, Sirt]; Asma Abufanas*, Hussameddin Badi*, Israa Benismai*, Hawa Obeid*, Abdulwahab Abdalei, Ahmed Abdulrahman, Aisha Swalem, Ebtisam Alzarouq, Amna Safar, Esra Shagroun, Boshra Hashem, Fatheia Elrishi, Fatima Abdulali, Habeeba Ahmed, Ibrahim Eltaib, Joma Elzoubia, Aisha Albarki, Hoda El Mugassabi, Fatima Abushaala, Amany Abuzaho, Nida Juha, Raneem Egzait, Sundes Shetwan, Alzahra Lemhaishi, Faisel Matoug [Misurata Central Hospital, Misurata]; Eman Abdulwahed*, Aamal Askar, Abir Ben Ashur, Adel Bezweek, Bushra Altughar, David Emhimmed, Donia Elferis, Laila Elgherwi, Enas Soula, Doaa Gidiem, Maren Grada, Khawla Derwish, Maram Alameen, Nassib Algatanesh, Ahlam Elkheshebi, Reem Ghmagh, Sharf Barka, Sultan Ahmeed, Sarah Aljamal, Zahra Alragig, Mohamed Addalla, Ahmed Atia, Atab Kharim, Fathia Mahmoud, Muhannud Binnawara, Entisar Alshareea [Tripoli Central Hospital, Tripoli]; Mohamed Alsori*, Aisha Alshawesh, Ghaliya Mohamed H Alrifae, Amira Ashour, Anwaar Abozid, Asil Omar Saleh Alflite, Anwar Mohamed, Jaber Arebi, Fatma Alagelli, Hana Yousef Gineeb, Rawia Ghmagh, Rihab Mohammed Bin Omar, Retaj Alaqoubi, Sara mohammed, Serien Hossain Bensalem, Tahani Elgadi, Wesam Sami, Yara Bariun, Abdulhadi Mohammed Alhadi Alhashimi, Dheba Almukhtar Abdulla, Heba Rhuma, Husam Enaami, Asraa Ali Alboueishi [Tripoli Medical Center/ Tripoli University Hospital, Tripoli]; Hayat Ben Hasan* [Zliten Medical Centre, Zliten]

#### Lithuania

Narimantas Samalavicius*, Vitalijus Eismontas, Jonas Jurgaitis, Oleg Aliosin, Vitalija Nutautiene [Klaipeda University Hospital]

#### Malaysia

Andee Dzulkarnaen Zakaria*, Anil Kumar Sree Kumar Pillai, Dinesh Kumar Vadioaloo, Mohamed Ashraf Mohamed Daud, Jien Yen Soh, Mohd Zaim Zakaria [School of Medical Sciences & Hospital USM, Universiti Sains Malaysia]; Siti Mayuha Rusli*, Nur Ayuni Khirul Ashar*, Zatul Akmar Ahmad*, Afiq Aizat Ramlee, Sharifah Nor Amirah Syed Abdul Latiff Alsagoff, Ahmad Anuar Sofian, Muhammad Badrul Hisyam Mohamad Jamil, Bahiyah Abdullah, Mohamad Faiz Noorman, Muhammad Fihmi Zainal Abidin, Mohamed Izzad Isahak, Siti Nasyirah Nisya Adnan, Zaidatul Husna Mohamad Noor, [Hospital Universiti Teknologi Mara (HUiTM)]

#### Mexico

Luis Adrian Alvarez-Lozada*, Alejandro Quiroga Garza, Andrea Aguilar Leal, Bernardo Alfonso Fernández Reyes, Ethel Valeria Orta Guerra, Francisco Javier Arrambide Garza, Héctor Erasmo Alcocer Mey, Jorge Arath Rosales Isais, Juventino Tadeo Guerrero Zertuche, Patricia Ludivina González García, Luis Antonio Heredia Sánchez, Marcela Patricia Flores Mercado, Oscar Alonso Verduzco Sierra, Pedro Emiliano Ramos Morales, Stephie Oyervides Fuentes, Víctor Manuel Peña Martínez [University Hospital Dr. Jose Eleuterio Gonzalez, Monterrey, Nuevo Leon]

#### New Zealand

Surya Singh*, Arwa Hadi, Christian Woodbridge, David Thornton-Hume, Jack Forsythe, Isini Dharmaratne, Vivian Pai, John Windsor, Kamran Zargar, Lucy Waldin, Lily Winthrop, Matias Alvarez, Meileen Huang, Matt Kumove, Marta Simonetti, Namisha Chand, Oliver Goldsmith, Oscar Guo, Paul Monk, Karen Zhou, Sai Harshitha Penneru, Shaamnil Prasad, Seifei Ren, Terrence Hill, Vyoma Mistry, Selena Sun [Nz, Auckland, Auckland City Hospital]; Ashley Pereira*, Scott Mclaughlin*, Andrew Stokes, Avinash Sathiyaseelan, Jeremy Rossaak, Janice Lim, Kenya Brooke, Liam Quinlan, Mark Pottier, Nayanika Podder, Puja Jinu, Shanay Ramphal, Wikus Vermeulen [Nz, Bay Of Plenty, Tauranga Hospital]; Fraser Jeffery, Ibrahim S. Al Busaidi, Janelle Divinagracia, William Ju, Yizhuo Liu, Tamara Glyn, Nasya Thompson* [Nz, Canterbury, Christchurch Hospital]; Vivien Graziadei*, Joshua Canton*, Joseph Furey*, Horim Choi, Grace Coomber, Tanya Divekar, Tessa English, Erin Gernhoefer, Tom Healy, Justin Chou, Dikshya Parajuli, Catherine Reed, Rod Studd, Anthony Lin [Nz, Capital And Coast, Wellington Hospital]; Cameron Wells*, Cindy Xu*, Arwa Hadi, Andrew Maccormick, Heejun Park, Athulya Rathnayake, Brittany Williams, Ashley Chan, Corinne Smith, Francesca Casciola, Jainey Bhikha, Jonathan Luo, Kevin Yi, Megan Singhal, Ria George, Rosie Luo, Taylor Frost [Nz, Counties Manukau, Middlemore]; Fatima Hakak*, Akhita George, Angela Carlos, Annie Ho, Connor Mcrae, Jonathan Lescheid, Jenny Soek, Andrew Pham, Sophie St Clair, Su-Ann Yee, Jennifer Lim [Nz, Lakes, Rotorua Hospital]; Taehoon Kim*, Anne Qi Chua, Christopher Harmston, Hamish Boyes, Holly Cook, Jamie Struthers, Jess Radovanovich, Nicholas Quek [Nz, Northland, Whangarei Base Hospital]; Chekodi Fearnley-Fitzgerald*, Deborah Wright, Kushan Ghandi, Natalie Matheson [Nz, Southern, Dunedin Hospital]; Matthew James McGuinness*, Brian Chen, Rebecca Indiana Douglas, Konrad Richter, Nisha Bianca Soliman, Scott Matthew Bolam, Vineeth Vimalan, William Currie [Nz, Southern, Invercargill (Kew) Hospital]; Mitchell Cuthbert*, Poppy Ross*, Amy Nicholson, Briar Garton, Emilie Agnew, Niamh Conlon, Nicholas Waaka, Ritwik Kejriwal, Sean Nguyen, Edmund Leung [Nz, Taranaki, New Plymouth Hospital]; Milidu Ratnayake*, Quintin Smith*, Nejo Joseph*, Bosco Yue, Calvin Fraser, Charles Lam, Ethan Figgitt, Gordon Liu, Kevin Tan, Ha Seong You, Helen Zheng, Jenny Luo, James Sharp, Kabir Khanna, Levi Simiona, Michel Luo, Milidu Ratnayake, Patrick Wong, Rebecca Luu, Rohit Paul, Shiva Nair, Shadie Asadyari-Lupo, Wing Hung, Geoffrey Ying [Nz, Waikato, Waikato Hospital]; Jess Ho*, Alan Wu, Eamon Walsh, Jouyee Lee, Jessie Liu, Sunny Yao, Omar Nosseir, Jennifer Dang, Simon Young, Sof’ya Zyul’korneeva, Theresa Boyd [Nz, Waitemata, North Shore Hospital]; Jess Ho*, Alan Wu, Sunny Yao, [Nz, Waitemata, Waitakere Hospital]

#### Nigeria

Abdullahi Musa Kirfi*, Adamu Bala Ningi, Mohammad Albuhari Garba, Makama Baje Salihu, Ohia Ernest Ukwuoma, Abdullahi Ibrahim, Isa Mienda Sajo, Muhammad Baffah Aminu, Liman Haruna Usman, Oloko Nasirudeen Lanre, Ibrahim Shaphat Shuaibu, Stephen Yusuf, Tiamiyu Ismail, Gabi Ibrahim Umar [Abubakar Tafawa Balewa University Teaching Hospital Bauchi, Bauchi]; Ademola Adeyeye*, Ehis Afeikhena, Favour Nnaji, Joy Agu, Temi Maxwell, Tosin Motajo, Karo Ifoto, Seubong Okon [Afe Babalola University Multisystem Hospital, Ado Ekiti]; Jerry Godfrey Makama*, Amina Abosede Mohammed-Durosinlorun, Bashiru Aminu, Polite Iwedike Onwuhafua, Caleb Mohammed, Lubabatu Abdulrasheed, Joel Amwe Adze, Khadijah Richifa Suleiman, Lydia Regina Airede, Mathew Chum Taingson, Stephen Bodam Bature, Stephen Akau Kache, Uchechukwu Ohijie Ogbonna [Barau Dikko Teaching Hospital, Kaduna]; Mohammed Bello Fufore*, Abdulkarim Iya, Adeshina A Ajulo, Ahmad Mahmud, Bilal Shuaibu Yahya, Farida Onimisi-Yusuf, Hope Isaac, Timothy Jawa, Fashe Joseph, Bemi Kala, Maisaratu A Bakari, David Wujika Ngwan, Abubakar Umar, Abraham L Filikus, Daniel Wycliff [Modibbo Adama University Teaching Hospital, Yola]; Abiodun Okunlola*, Olukayode Abiola, Adebayo Adeniyi, Olabisi Adeyemo, Babatunde Awoyinka, Olakunle Babalola, Adewumi Bakare, Taiwo Buari, Cecilia Okunlola, Gbadebo Adeleye, Adedayo Salawu, Henry Abiyere, Adetolu Ogidi, Tesleem Orewole [Federal Teaching Hospital, Ido Ekiti]; Habiba Ibrahim Abdullahi*, Godwin Akaba, Arome Achem, Asi-oqua Bassey, Emeka Ayogu, Bilal Sulaiman, Dennis Anthony Isah, Chukwunonso Nnamdi Akpamgbo, Felicia Asudo, Nathaniel Adewole, Omachoko Oguche, Peter Ejembi, Samuel Ali Sani, Paul Chimezie Andrew, AliyuYabagi Isah, Bolarinwa Eniola, Zumnan Songden, Teddy Agida, Terkaa Atim [University Of Abuja Teaching Hospital, Gwagwalada]; Taofiq Olayinka Mohammed*, Hadijat Olaide Raji*, Femi Ibiyemi, Hafeez Salawu, Olushola Fasiku, Remi Sanyaolu Solagbade, Mariam Motunrayo Shiru, Gbadebo Hakeem Ibraheem, Justina Oruade, Grace Ezeoke [University Of Ilorin Teaching Hospital, Ilorin]

#### Pakistan

Tabish Chawla*, Aliya Begum Aziz, Anoosha Marium, Ayesha Akbar Waheed, Faiqa Binte Aamir, Faiza Qureshi, M Hammad Ather, Iqra Fatima Munawar Ali, Izza Tahir, Maha Ghulam Akbar, Ronika Devi Ukrani, Sajjan Raja, Sehar Salim Virani, Shahryar Noordin, Saif Ur Rehman, Shalni Golani, Syed Roohan Aamir, Syed Musa Mufarrih, Usama Waqar, Maliha Taufiq [Aga Khan University Hospital]; Ahmed Siddique Ammar*, Adya Ejaz*, Albash Sarwar, Ahmed Usman Khalid, Shehrbano Khattak [Bahria International Hospital Lahore]; Aliza Imran, Omer Bin Khalid, Urauba Kaleem, Urwah Muneer, Yumna Kashaf [Creek General Hospital]; Fatima Zafar*, Adil Zaheer, Muhammad Ali, Amna Shafaat, Arisha Qazi, Asjad Imran,Mahnoor Tariq, Muhammad Nadeem Aslam, Shehroz Ali, Tabish Atiq, Tayyiba Wasim, Daniyal Babar, Ahmad Zain, Muhammad Ibtisam [Services Hospital Lahore]; Uzair Ahmed, Syed Talha Bin Aqeel, Muhammad Muhib, Muhammad Anas Abbal, Nasar Ahmad Khan, Imran Javed [United Hospital]

#### Palestine

Layth Alkaraja*, Dana Amro, Ghaida Manasrah, Ibraheem Hammouri, Ihab Abu Hilail, Jihad Zalloum, Laith Alamlih, Mahmoud Nasereddin, Munia Rajabi, Sa’ed Shalalfeh, Zeinab Natsheh [Hebron Government Hospital, Hebron]; Khamis Elessi*, Mustafa Abu Jayyab*, Mohammed Astal, Mosheer Al-Dahdouh [Nasser Medical Complex, Gaza]; Alaa Eddin Salameh*, Alaa Ayyad, Nimatee Dawod, Hamza Alsaid, Iyas Matar, Majd Hassan, Mohammed Bakeer, Mohammad Malasah, Shehab Abuhashem, Mohammed Salem, [Palestine Medical Complex, Ramallah]

#### Romania

Sorinel Lunca*, Mihail Gabriel Dimofte, Stefan Morarasu, Ana Maria Musina, Cristian Ene Roata, Natalia Velenciuc [Regional Institute Of Oncology Iasi, Iasi]

#### Russia

Aleksandr Butyrskii*, Maxim Bozhko, Amet Ametov [Emergency Municipal Hospital No.6]

#### Saudi Arabia

Sharfuddin Chowdhury*, Doaa Bagazi [King Saud Medical City, Riyadh]

#### Spain

Julio Domenech*, Alejandro Rosello-Añon, Ana Monis, Caterina Chiappe, Beatriz Cuneo, Pablo Clemente-Navarro, Jorge Febre, Jorge Sanz-Romera, Marcos Lopez-Vega, Ignacio Miranda, Rocio Valverde-Vazquez, Sara Garcia, Maria Jose Sanguesa, Zutoia Balciscueta [Hospital Arnau De Vilanova]; Enrique Ruiz*, Eduardo Marco, Elena Talavera, Joan Farre, Loreto Bacariza, Mireia Duart, Violeta Ureña, Xenia Carre [Hospital Sant Joan Reus]

#### Sudan

Hytham K. S. Hamid*, Montasir A. Abd-Albain, Sami Galal-Eldin [Al-Moalem Medical City]; Monira Sarih*, Eithar Adam, Samir Ismail, Malaz Azhari, Tawfieg Hassan [Alandalus Clinic, Elduiem]; Mohamed Salaheldein*, Zainab Abdalla, Wahiba Ahmed [Bashair Teaching Hospital, Khartoum]; Monzer Abdulatif Mohamed Alhassan*, Hozifa Mohamed Abdalla Suliman, Hozifa Mohamed Bdalla Suliman [Education Karima Hospital, Merowe]; Rogia Ahmed Abdalla Ahmed*, Enas Mohammedtom Abdulhameed Babekir, Munya Ali Talab Khairy, Maha Mukhtar Ahmed Mukhtar, Rzan Ali Hamedelneel Ali, Yasir Babkir Ali Al-Shambaty [Elduiem Teaching Hospital, Elduiem]; Fatima Imad Yousif*, Hawa Mohammed Hassan Mohammed, Lana Osher, Lana Osher, Menhag Abdelbast, Mohamed Yassin, Noon Moawia, Rowa Abdalsadeg [Gadarif Teaching Hospital, Gadarif City]; Abrar Husein, Baraa Elhassan, Alnazeer Y. Abdelbagi, Mohammed A. Adam, Eithar M. Ali, Ibrahim A.b. Mohammed, Maab Mohamed, Mohamed Abdulaziz, Mazin Akasha, Muaz Hassan, Nadir Hilal, Noon Abdalla Abdelrahman Mohamed, Noora Abubaker, Omeralfarouk Mohammed, Shakir Mohamed, Walaa Osman, Fatima Mustafa, Alaa A Salih [Ibn-Sina Hospital, Khartoum]; Doua Ali*, Doha Mohammed Ahmed Almakki, Hanan Elnour Mohamed, Abdelhadi Elmubark, Mohamed Hassan, Ammar Alnour, Amna Elaagib, Ayman Abdelrahman, Mubarak Abdelkhalig, Khalid Nour Eldaim, Afra Babiker, Entisar Ahmed, Maab Ali, Eman Hussain, Mansour Wedatalla, Alaaaldeen Ahmed, Alla Aldeen Hamza, Mohab Mohammed, Omer Osman, Reham Ibrahim, Rihab Ahmed, Ruaa Ahmed, Ruaa Yasir, Safaa Awadallah, Sara Mohmmed, Suhaib Hassan [Ibrahim Malik Teaching Hospital, Khartoum]; Walid Shaban*, Aisha Hussein, Reem Rafea, Ahmed Abdalla, Abdalla Ahmed, Khalid Mohamed, Mansour Mohammed, Mohamed Altahir, Mohammed Adam, Omer Mohamed, Walaa Abdullah [Khartoum North Teaching Hospital (Bahri Hospital)]; Hammad Fadlalmola*, Ahmed Yassir Abdalla, Ahmed Ali Omer, Ahmed Alfatih Mustafa, Rawan Elhadi, Essam Eldien Abuobaida Banaga, Fatima Osman, Mohamed Abdalla, Hala Abdelhalim Mohamed Taha, Noon Ezzeldien Abdalmahmoud, Rofuida Hussien Nafie, Sami Jamal, Sharwany Ahmed [National Ribat University Hospital, Khartoum]

#### Syria

Rawan Alsheikh Ali*, Abdallah Aladna, Abdullah Aljoumaa, Hamdi Nawfal, Salma Jamali, Fatima Khouja, Ammar Niazi, Toka Al Rawashdeh [Aleppo University Hospital, Aleppo]

#### Tunisia

Nahla Kechiche*, Mouna Gara, Mouna Nasr, Marwen Baccar, Oumayma Benamor, Sawssen Chakroun [University Hospital Fattouma Bourguiba, University Of Monastir]

#### Turkey

Ahmet Necati Sanli*, Ahmet Yildiz, Mehmet Ali Demirkiran, Yildiz Buyukdereli Atadag, Yusuf Iskender Tandogan [Abdulkadir Yuksel State Hospital]; Esin Ozkan*, Yıldırım Ozer, Esin Ozkan, Muhammed Miran Oncel, Senad Kalkan [Bezmialem Vakif University, Faculty Of Medicine, Istanbul]; Tolga Gover*, Berke Manoglu, Ilayda Oksak, Ipek Kurt, Kerem Rifaioglu, Selman Sokmen, Tayfun Bisgin, Yasemin Yildirim, Abdil Yetkin Keskin [Dokuz Eylul Univ. Hospital, Izmir]; Tugce Dogan*, Berfin İlgaz Sahin, Cemil Aydin, Duygu Ece Benek, Hale Nur Tiras, Mert Arslangilay, Mert Aslangilay, Muhammet Yaytokgil, Mehmet Ali Capar, Yasemin Yazgan [Hitit University Faculty Of Medicine Çorum Research And Training Hospital]; Sebnem Bektas*, Ahmet Can Alagoz, Alara Ece Dagsali, Aylin Izgis, Kadir Uzel, Mustafa Soytas, Niyazi Cakir, Abdullah Emre Askin, Ibrahim Azboy, Kubilay Sabuncu, Merve Aslan, Melek Sahin, Mustafa Oncel, Nuri Okkabaz, Ramazan Kemal Sivrikaya, Alparslan Saylar, Dr. Alparslan Saylar, Meltem Yasar [Istanbul Medipol University Hospital, Istanbul]; Ergin Erginoz*, Haktan Ovul Bozkir, Kagan Zengin, Mehmet Faik Ozcelik, Server Sezgin Uludag, Zeynep Ozdemir [Istanbul University Cerrahpasa - Cerrahpasa School Of Medicine]; Osman Sibic*, Hatice Telci, Mehmet Abdussamet Bozkurt, Yasin Kara [Kanuni Sultan Suleyman Training And Research Hospital, Istanbul]; Mustafa Deniz Tepe*, Adnan Gündoğdu, Bilge Akın, Dilan Pehlivan, Ali Guner, Duygu Baysallar, Berkay Yıldız, Hale Cepe, Murat Emre Reis, Ayse Nilufer Yuzgec, Nurtac Kıralı, Taha Anıl Kodalak, Mehmet Ulusahin [Karadeniz Technical University Farabi Hospital, Trabzon]; Kamar Selim*, Ahmet Kale, Mehmet Emre Gecici, Melis Ozbilen [Kartal Dr. Lutfi Kirdar Training And Research Hospital, Istanbul]; Zeynep Düzyol*, Aylin Gemici, Elzem Korkmaz, Eminenur Şen, Muhammed Enes Taşcı, Elifsu Camkıran, Güşta Elieyioğlu, İkbal Kayabaş, Tevfik Kıvılcım Uprak, Canan Aral, Ayten Saraçoğlu, Mustafa Ümit Uğurlu, Zeynep Hazal Baltacı [Marmara University, School Of Medicine, Istanbul]; Ege Nur Akkaya*, Cem Fergar, Elif Zeynep Tabak, Guldane Zehra Kocyigit, Ilgaz Kayilioglu [Mugla Training And Research Hospital, Mugla]; Süleyman Polat*, Eli□f Çolak, Mehmet Emin Kara, Mert Candan, Mustafa Safa Uyanık, Ahmet Can Sarı [Samsun Training And Research Hospital, Samsun]; Attila Ulkucu*, Alperen Taha Certel, Arzu Dindar, Beyza Durdu, Cigdem Bayram, Eslem Kaya, Hakan Akdere, Ibrahim Ethem Cakcak, Ikranur Yavuz, Mert Omur, Mirac Ajredini, Erhan Onur Aydoğdu, Eylül Şenödeyici [Trakya University Faculty Of Medicine]; Ulku Ceren Koksoy*, Baturay Kansu Kazbek, Deniz Serim Korkmaz, Dogancan Yavuz, Hakan Yilmaz, Zeynep Sahan Cetınkaya, Elif Durmus, Filiz Tuzuner, Furkan Hokelekli, Mucahid Mutlu, Seyma Orcan Akbuz, Ziya Can Kus, Ziya Can Kus [Ufuk Üni□versi□tesi□ Tip Fakültesi□ Dr.Ri□dvan Ege Sağlik Araştirma Uygulama Merkezi□ Hastanesi□, Ankara]

#### United States of America

Michael Farrell*, Alayna Craig-Lucas, Matthew Painter, [Lehigh Valley Health Network]; Ashley Titan*, Aditya Narayan, Bunmi Fariyike, Lisa Knowlton, Tiffany Yue [Stanford Health Care, Palo Alto, California]; Emily Benham*, Abdelrahman Nimeri, Hope Werenski, Nicole Kaiser, Caroline Reinke [Atrium Health]

*Local lead

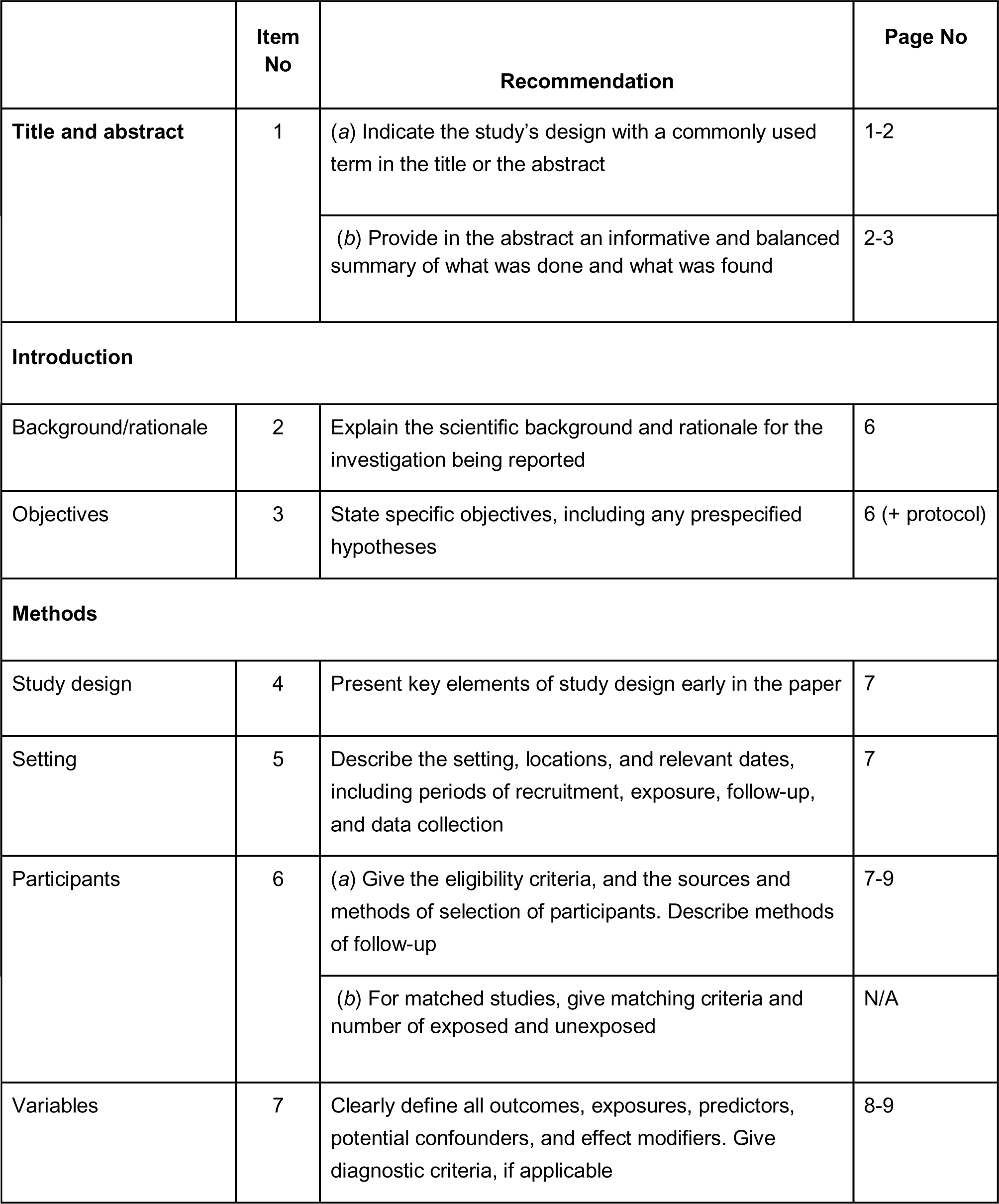
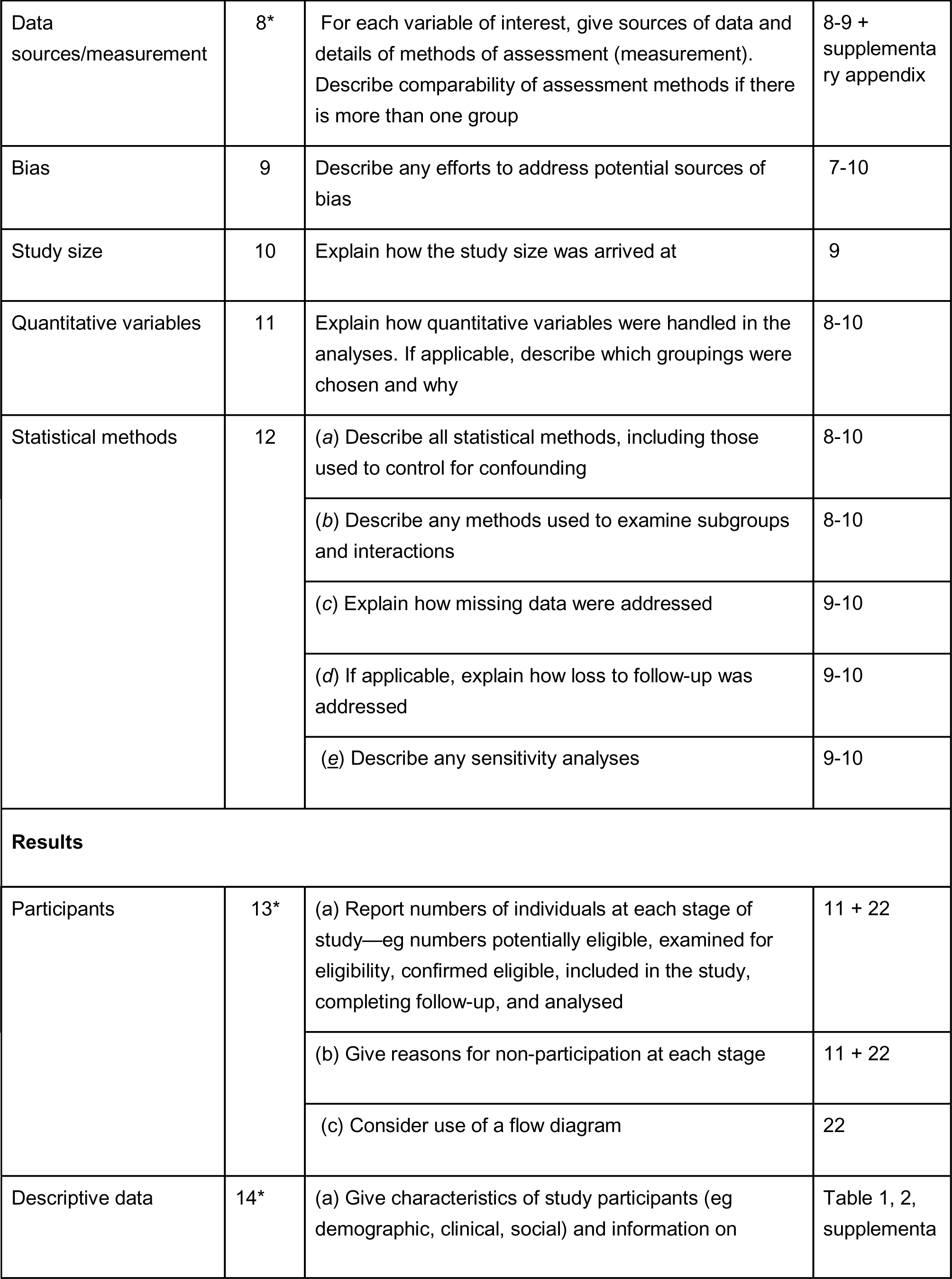
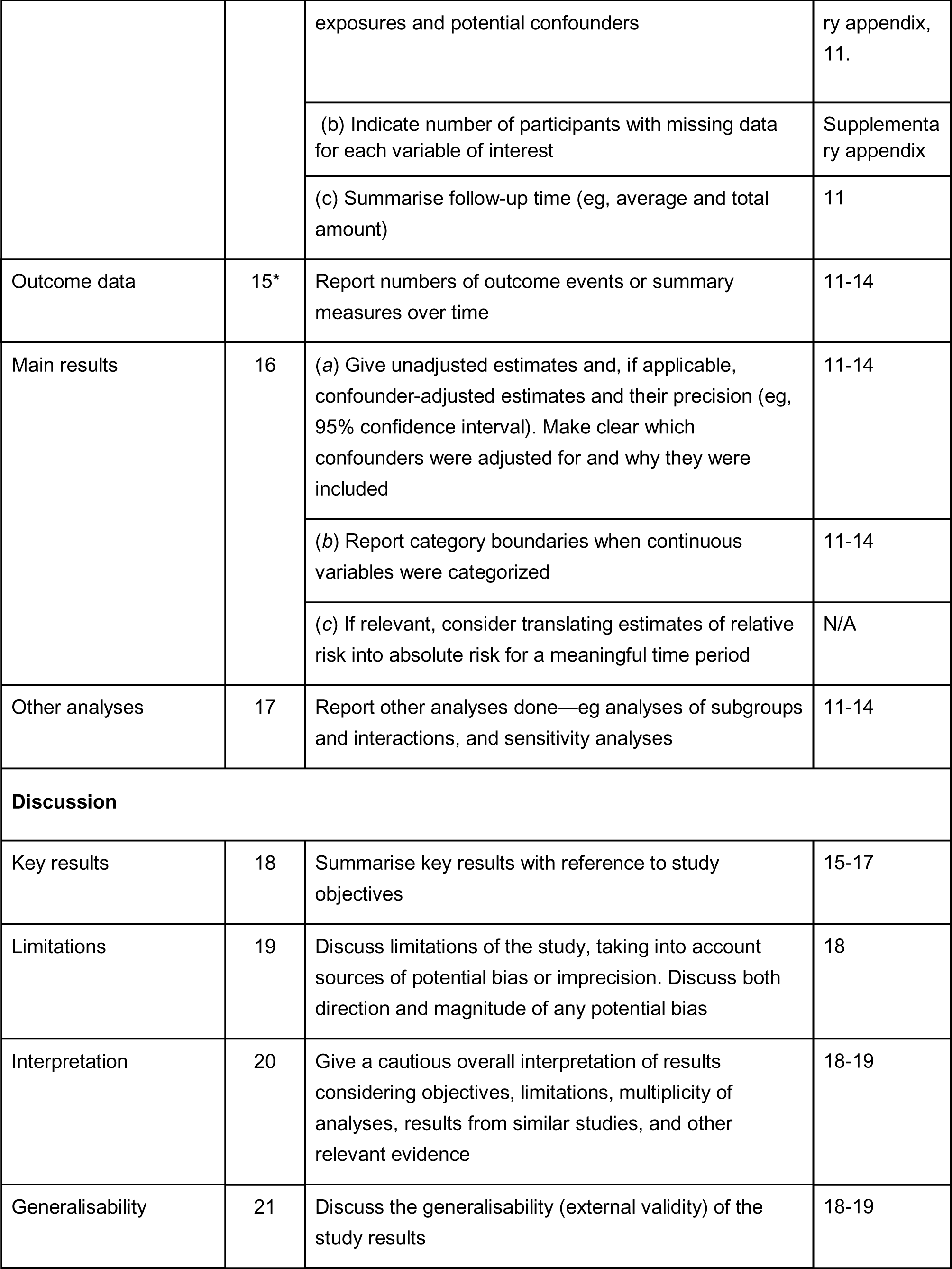
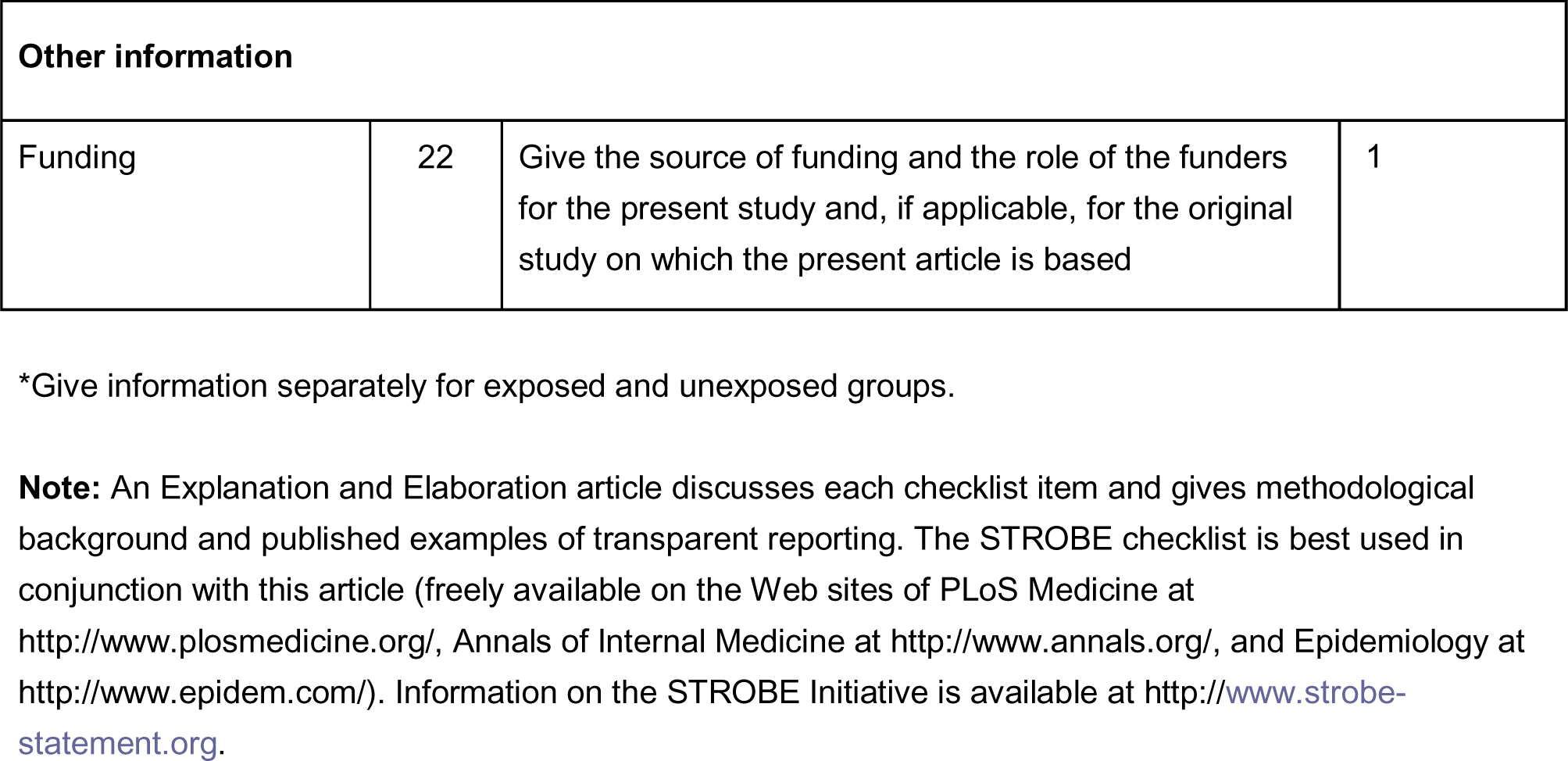
**STROBE Statement**—Checklist of items that should be included in reports of *cohort studies*

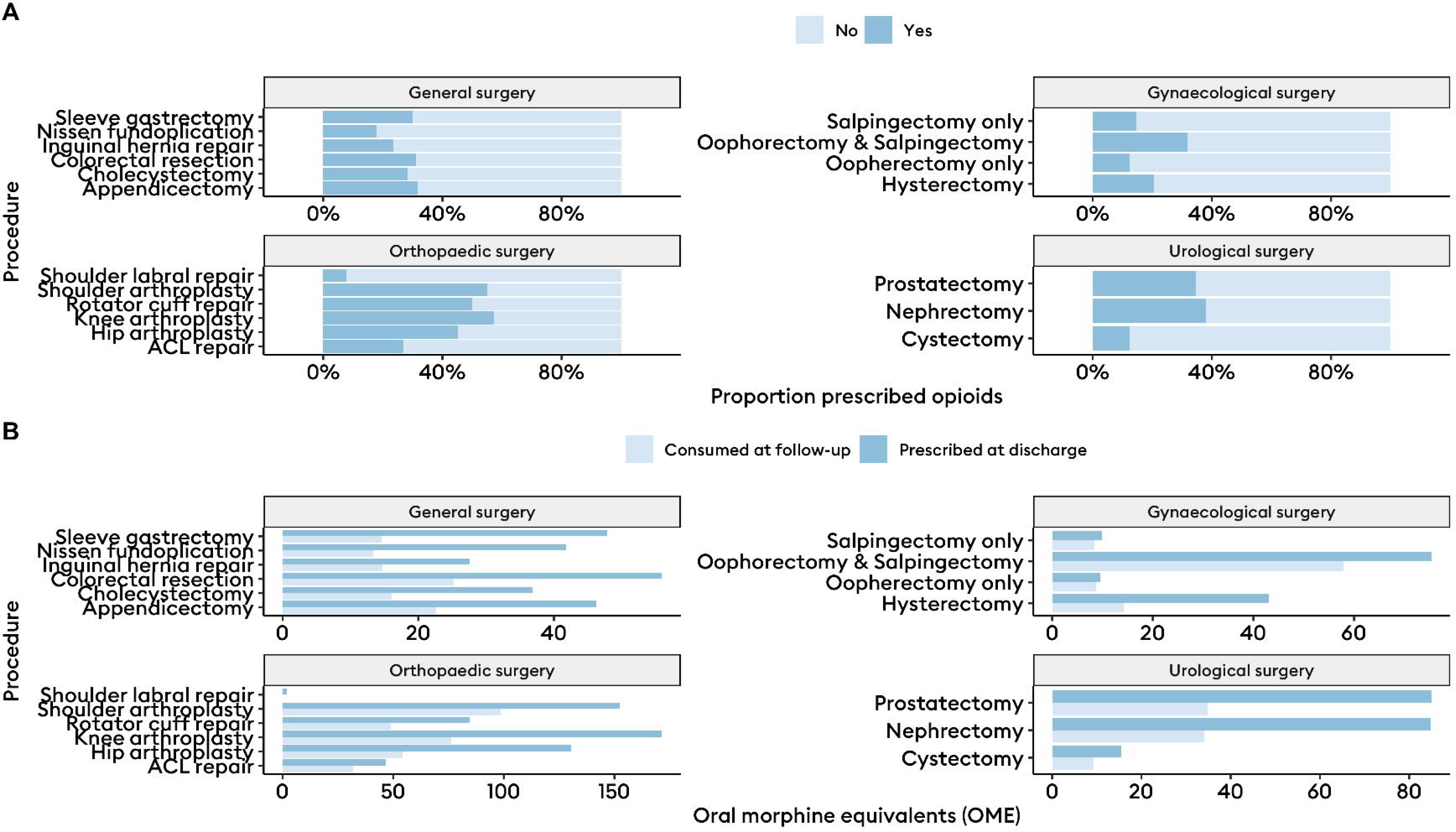

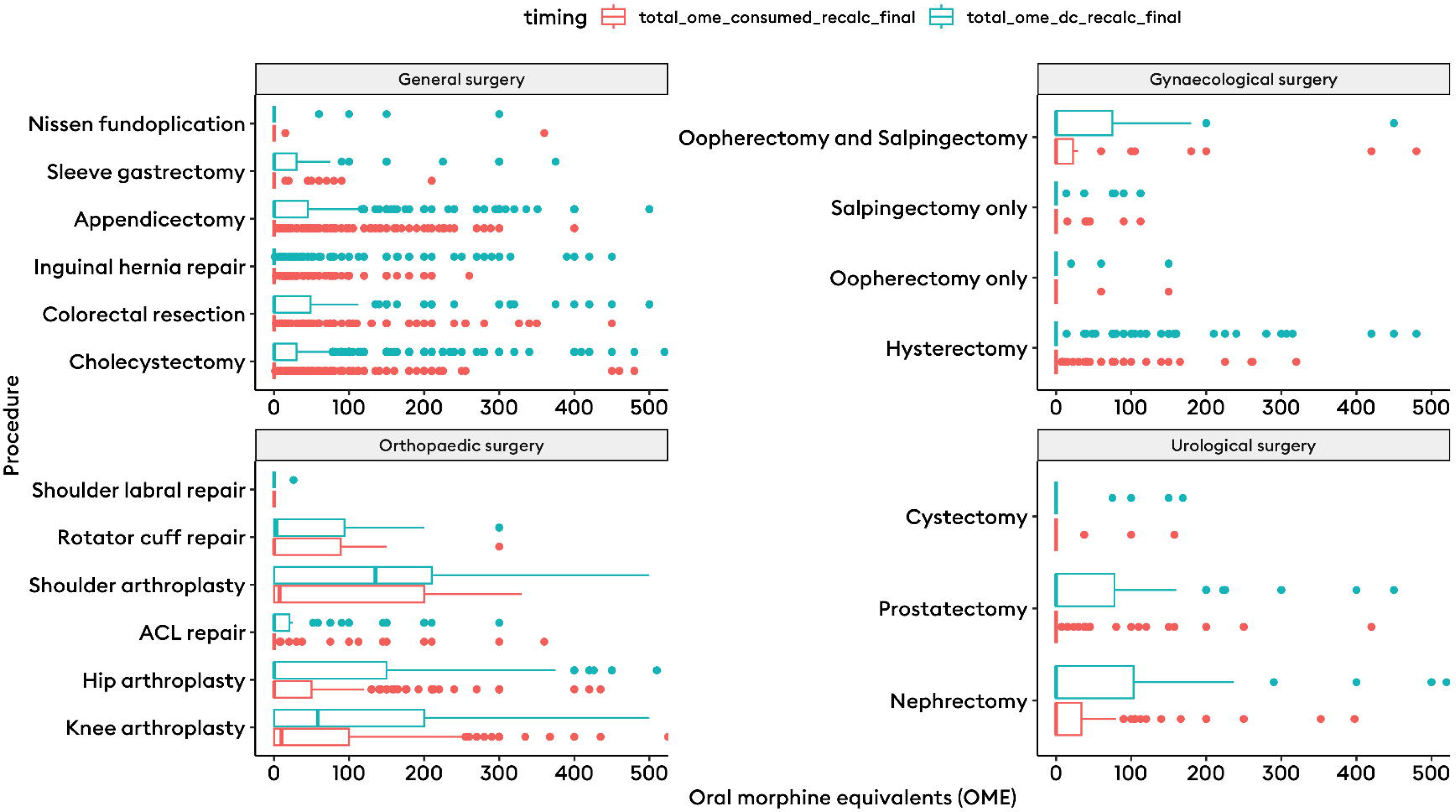

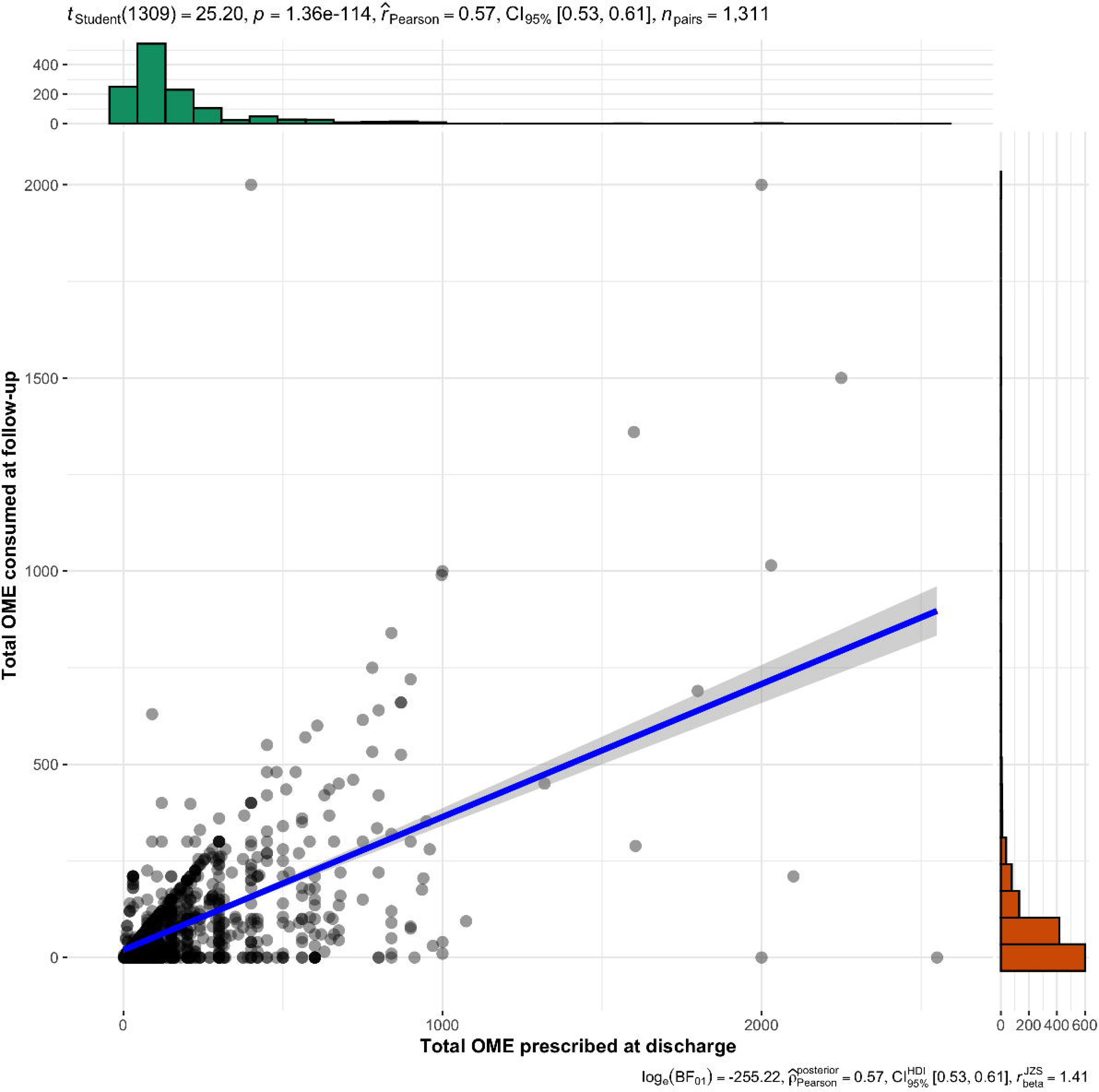

